# GWAS on short tandem repeats identifies novel genetic mechanisms in Alzheimer’s disease

**DOI:** 10.1101/2025.03.13.25323833

**Authors:** David Gmelin, Olena Ohlei, M. Muaaz Aslam, Laura Parkkinen, Kristina Mullin, Dmitry Prokopenko, Christina Lill, Rudolph E. Tanzi, Valerija Dobricic, Lars Bertram

## Abstract

Genome-wide association studies (GWASs) are typically based on the analysis of single nucleotide polymorphisms (SNPs) and often exclude more complex genetic variants, such as short tandem repeats (STRs). Here, we report the results of GWAS analyses systematically assessing the role of STRs, both imputed and directly genotyped by whole genome sequencing (WGS), on risk for Alzheimer’s disease (AD) in a large collection of ∼330,000 individuals (3,287 AD cases; 47,048 AD-by-proxy cases, 283,111 controls) from the UK biobank. Using imputed (or WGS-derived) STR genotype data, we identified 14 (WGS: one) independent loci showing evidence for genome-wide significant association with AD risk.

While most identified loci had already been highlighted by SNP-based GWAS, we detected new STR-based signals near the genes *SNX32* (chr. 11q13) and *WBS1* (chr. 17q11). In addition, we delineated several other loci where STRs (and not SNPs) either represent the lead signal (*ABCA7*) or make substantial contributions to the SNP-driven associations (*HLA-DRB1, MINDY/ADAM10*, and *APOE*). Heritability analyses estimated that STRs account for at least 3% of the total phenotypic variance of AD in this dataset. Aligning our top STRs with DNA methylation and transcriptome profiles from human brain samples suggests that several STRs may unfold their effects by impacting gene expression. Future work needs to confirm our results and delineate the likely considerable role that STRs play in the genetic makeup of AD.

## Introduction

Alzheimer’s disease (AD) is a genetically complex neurodegenerative disorder. For early-onset familial AD (EOFAD), rare, disease-causing mutations have been identified in three genes *(APP*, *PSEN1*, *PSEN2*), typically leading to an onset before 65 years^1^. However, the vast majority of AD cases has a later disease onset (commonly referred to as late-onset AD [LOAD]) genetically governed by a polygenic risk architecture^2^. For this latter form, genome-wide association studies (GWAS) have uncovered common variants (typically single nucleotide polymorphisms [SNPs]) at more than 70 loci significantly modifying disease risk^3^. While GWAS have substantially enhanced our understanding of AD genetics, a large portion of the phenotypic variance remains unexplained by the hitherto known loci^4^, a situation often referred to as “missing heritability”^5^.

A sizeable portion of this missing heritability may be elicited by the effects of other types of genetic variants, such as tandem repeat (TR) polymorphisms. TRs represent repetitive elements of the genome consisting of units of specific nucleotide sequences (“repeats”) occurring adjacent to each other in a head-to-tail fashion (“tandem”). Short tandem repeats (STRs) are a subcategory of TRs where the repeat unit size is between 1 and 6 base pairs(bp)^6^. It is well established that STRs may represent a direct cause of disease through expansions of the TR sequence (e.g., STR expansions causing Huntington’s disease or fragile X syndrome^7^). Typically, these TR expansions are very rare and function like monogenic disease-causing mutations. Notwithstanding, the length of most STRs is highly variable and these length polymorphisms have been associated with gene expression^8^, mRNA splicing^9^, DNA methylation^10^, as well as disease risk^11,12^ emphasizing their role in shaping diverse human traits.

Despite their importance, a systematic analysis of STR effects on a genome-wide scale was hindered by technological limitations until recently. This has now changed with the advent of cost-efficient whole genome sequencing (WGS) approaches and the development of TR-aware computational tools^13,14^. Together, these developments have led to the creation of WGS-derived STR reference panels that allow to impute STR polymorphisms from SNP genotyping data^15,16^. Despite these developments, there are still comparatively few studies that have interrogated the role of STRs in complex traits at a genome-wide scale. These have either used imputed STRs^11,12,17,18^ or STRs called directly from WGS data^19,20^. Collectively, the (sparse) literature available on the topic to date suggests that 5-10% of complex trait GWAS signals may be elicited by STRs rather than SNPs^11,18^. For AD, we are only aware of one study on the topic^19^. Perhaps owing to its comparatively small sample size (n∼3,000), the only locus identified to show significant STR-related effects in that study was *APOE*^19^. In summary, STR-based GWAS are still rare for complex traits, including AD. The few studies that have been published to date concluded that STRs likely make an important and hitherto underappreciated contribution to the genetic architecture of complex traits.

Here, we set out to thoroughly and systematically assess the potential role of STR polymorphisms in contributing to AD risk. To this end, we performed STR-based GWAS analyses using both imputed and WGS-derived STR data in 333,446 (3,287 AD cases; 47,048 AD-by-proxy cases) and 107,289 (1,026 AD cases, 15,089 AD-by-proxy cases) samples of the UK biobank (UKB), respectively. The interpretation of our primary findings is buttressed by SNP-based GWAS analyses and extensive *in silico* fine-mapping to disentangle the respective roles of SNPs vs STRs in contributing to AD risk. Overall, we identify 254 imputed STRs across 14 independent loci and one WGS-derived STR to show genome-wide significant association with AD risk in this dataset suggesting novel genetic mechanisms in AD.

## Results

### Genome-wide association analyses on imputed STRs

After quality control (QC), we were able to include a total of 333,446 genetically unrelated UKB participants (of different ancestries; Methods), comprising 3,287 clinically diagnosed AD cases, 47,048 AD-by-proxy cases, and 283,111 controls (Supplementary Figure 1, Supplementary Table 1). For discovery, we performed GWAS analyses on 295,551 individuals self-reported as “White-British” (discovery cohort: 2,947 AD cases, 42,923 AD-by-proxy, and 249,681 controls; Supplementary Table 1) and 3,026,404 imputed STR variants that passed QC (Methods). These analyses resulted in 254 variants at 14 independent loci that passed our genome-wide significance threshold after multiple testing correction (p<1.49E-07; Methods). In addition, there were 228 variants across 21 independent loci showing at least genome-wide suggestive (alpha=1.0E-05) evidence of association (Figure 1, Supplementary Table 2). Comparison of observed vs. expected genome-wide test statistics revealed a λ of 1.056 suggesting that our STR-based GWAS results are not affected by undue inflation (Supplementary Figure 2A).

**Figure 1.**
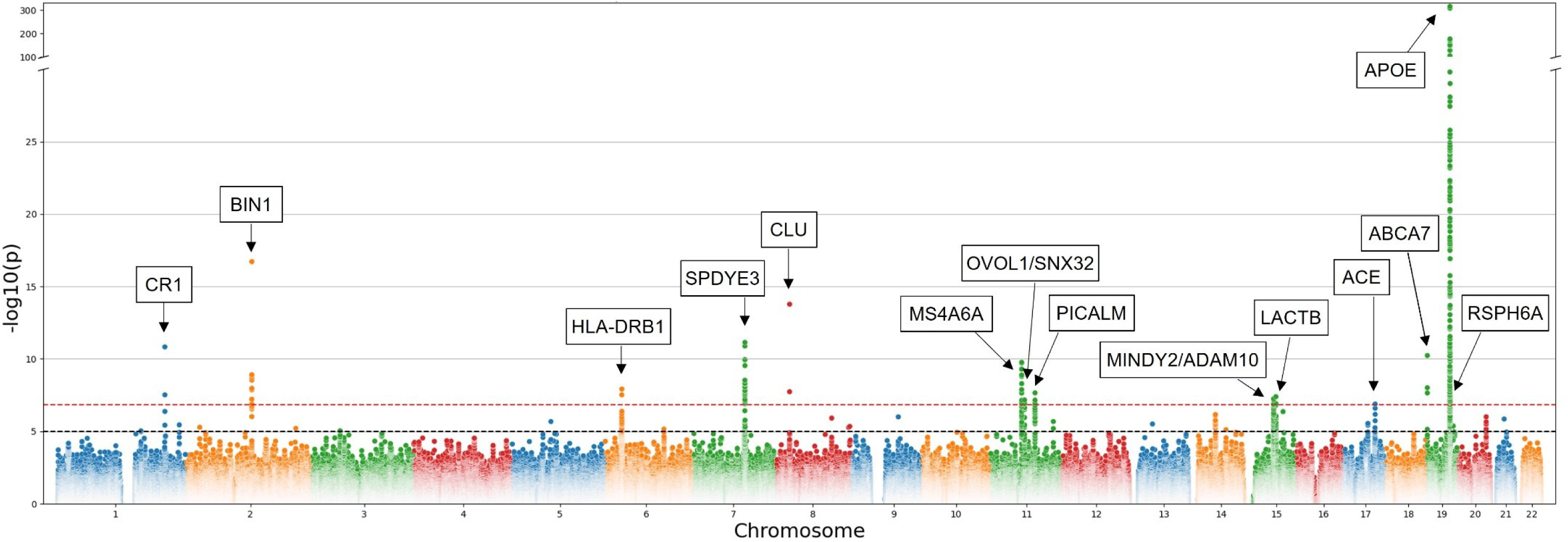
Manhattan plot showing results of GWAS using linear regression for AD and AD-by-proxy status and imputed STRs on “White-British” cohort. In each box, the nearest protein-coding gene according to GENCODE V47 is annotated. Horizontal red dashed line indicates the genome-wide significance threshold of 1.49E-07, whereas the black dashed line indicates the suggestive significance threshold of 1.00E-05. Note that y-axis is discontinuous and capped at 300. See Supplementary Table 3 for more details on these results.

Most of the detected genome-wide significant STR associations (n=153 unique STRs) were within the *APOE* region on chromosome 19q13.32, a well-established and likely SNP-based risk factor for AD^21,22^. The remaining 76 STRs mapped to 13 independent loci outside the *APOE* region, most often to the vicinity (i.e. 1.3 – 138.5 kb distance) of known AD GWAS signals (based on reference 3). Only one STR signal (i.e. on chr11q13.1) mapped into an intergenic region outside known AD GWAS signals (nearest genes *OVOL1* and *SNX32*; Figure 1). In eight of the 13 non-*APOE* loci, STRs were located within the open reading frame (ORF) of protein coding genes (all intronic), while five were between ∼5 and 27kb up-/down-stream from the nearest protein coding gene (Supplementary Table 3).

With respect to the 228 suggestively significant STRs, the majority (n=178, across 21 independent loci) were located outside the *APOE* region (Supplementary Table 4). Thirteen of these peaks are located more than 500kb from any known AD locus and, hence, may at least partially represent novel AD risk signals. Among these 13, eight are within the ORF (all intronic) while the remaining five STRs mapped between ∼2 and 310 kb up-/down-stream from the closest protein coding gene.

### Independent replication of primary STR-based GWAS results

To assess the validity of the findings in the discovery dataset, we generated analogous test statistics for all genome-wide significant STRs in an independent subset of white UKB individuals (“Other-White”; n=20,840; 185 AD cases, 2803 AD-by-proxy cases, and 17852 controls; Supplementary Table 1). These analyses revealed that six out of the originally identified 14 significant STRs showed direct evidence for replication (i.e. same effect direction and p<0.05; Supplementary Table 5). Of the remaining eight STRs, all but one showed the same direction of effect. Furthermore, meta-analyses across the discovery and “Other-White” subsets showed stronger associations (by p-value) for six out of these seven loci (Supplementary Table 5). Thus, 12 out of 14 identified genome-wide significant STR loci (i.e. all but *MS4A6A* and *OVOL1/SNX32*) showed either direct or indirect evidence for replication in “white” individuals independent of the discovery dataset. In addition, there was also support for many STR loci in GWAS results from the other descent groups (incl. *OVOL1/SNX32*; Supplementary Table 5). However, we note that the sample sizes from these non-white subsets were collectively small with AD numbers ranging from 93 to 422 (Supplementary Table 1), so replication evidence cannot be reliably interpreted in these data. Notwithstanding, the independent assessments in the “Other-White” subset of UKB revealed that most of the STR effects uncovered in the discovery GWAS analyses are stable and reproducible.

### Discerning the drivers of STR-based GWAS signals

Given that the STR imputation protocol used here relies on SNP genotypes, it can be expected that some of the imputed STR GWAS signals are actually driven by SNPs at the same locus. To assess this hypothesis, we performed a range of analyses aimed at discerning SNP from STR effects (Methods). As can be seen on the Manhattan plots from both analyses, the vast majority of genome-wide significant association peaks overlapped within a ±250kb window (Supplementary Figure 3). However, fine-mapping using GCTA-COJO nominated the STR in *ABCA7* as the driver of the association signal in this region.

Next, we performed a range of analyses using two conditioning paradigms: i) adjusting any of the 14 potential STR signal for SNP effects, and ii) vice versa (Methods and Supplementary Table 6). While in the first paradigm the original STR signal dropped above the suggestive threshold (alpha=1.0E-05) in all instances (Supplementary Table 6, Supplementary Figures 4 - 18), we note that two STRs (near *MINDY2*/*ADAM10* and near *ABCA7*) showed a residual, almost-suggestive association after conditioning (p=3.36E-05 and p=4.23E-05, respectively; Supplementary Table 6). This indicates that STRs may substantially contribute to the known genetic association signals in these regions. In addition to observing diminishing STR signals after adjusting for the effects of associated SNPs, we also observed the opposite, i.e., that certain STRs showed *stronger* evidence for association after conditioning. This was the case for the *HLA-DRB1* (six STRs) and *ABCA7* (one STR) loci (Supplementary Table 7). A similar situation was encountered in *APOE*, where one STR (chr19:44910674) showed a change in p-value from merely nominally significant (p=9.33E-03) to genome-wide significant (p=4.67E-23) (Supplementary Table 8) after conditioning on rs429358 (“e4-allele” in *APOE*).

This increase in statistical support was at least partially due to the effect of the “e2-allele” in *APOE* (rs7412), which is in pronounced LD with the associated STR allele (r2=0.71).

Interestingly, the situation of attenuated association evidence looked similar when applying the reciprocal paradigm (Supplementary Table 6, Supplementary Figures. 5 - 18). Except for *APOE* and two additional SNPs, i.e. rs6733839 within the *BIN1* region on chromosome 2 and rs117618017 within the *APH1B* region on chromosome 15, all SNP signals dropped above suggestive significance after adjusting for STR effects, possibly indicating that STRs also contribute to leading SNP-signals. We note that a clear interpretation of these analyses is aggravated by the fact that, by design, SNP and STR genotypes are highly correlated in this arm of our study (in contrast to analyses using WGS-derived STRs, see below). For suggestive STRs, these analyses resulted largely in a similar picture with a tendency towards more STRs “surviving” the SNP adjustment than for the genome-wide significant loci (Supplementary Table 9).

### Heritability estimates for STRs compared to SNPs

To estimate the contribution of STRs to the genetic architecture of AD beyond loci showing genome-wide significant evidence of association we computed genome-wide heritability estimates using the GCTA-LDMS method. Using a population prevalence of AD of 0.05^23^ and MAF>0.01, this resulted in a SNP-based heritability of 36.45%, which is in good agreement with recent estimates (31%) from other studies using GCTA for heritability estimation^24,25^.

Equivalent heritability computations on STR genotypes yielded an estimate of 31.16%. The estimate including both SNP and STRs combined maximized at 37.54%. Thus, STRs contribute minimally ∼3% of the total AD heritability (i.e. (h(SNP+STR)-h(SNP))/h(SNP+STR)), an estimate slightly smaller than for other complex traits ^11,18^. Notably, the estimated contribution of STRs to total AD heritability remained stable at ∼3% even when alternative prevalence values were used (here: 0.1, 0.2, 0.3). Notwithstanding, given the extensive correlation structure between SNPs and STRs in this dataset, it appears possible that some genuine STR effects are masked by SNP genotypes. Hence, we consider ∼3% to represent the lower bound of genetic AD heritability contributed by STRs.

### Genome-wide association analyses on WGS-derived STRs

To compare GWAS results from imputed vs. WGS-derived STR data, we utilized STR genotypes generated in the study by Halldorsson et al.^26^. After rigorous sample-and variant QC steps (Methods), a total of 95,201 UKB participants of self-reported “White-British” ancestry (henceforth referred to as “Halldorsson-White-British”) were available for GWAS (incl. 928 AD and 13759 AD-by-proxy cases; Supplementary Table 10). On the variant level, a total of 1,205,675 WGS-derived STRs were available for analysis. As expected based on the much smaller sample size, the number of signals was substantially reduced compared to the GWAS on imputed STRs (Figure 2, Supplementary Table 11). While 72 signals passed the genome-wide significance threshold (alpha=1.01E-07), only one of these (chr17:27264667 near *WSB1*) was located outside *APOE* (rs429358 ±0.5Mb). In addition, a total of 60 WGS-derived STRs across 19 loci showed suggestive evidence of association (25 STRs at 18 loci outside *APOE*).

**Figure 2.**
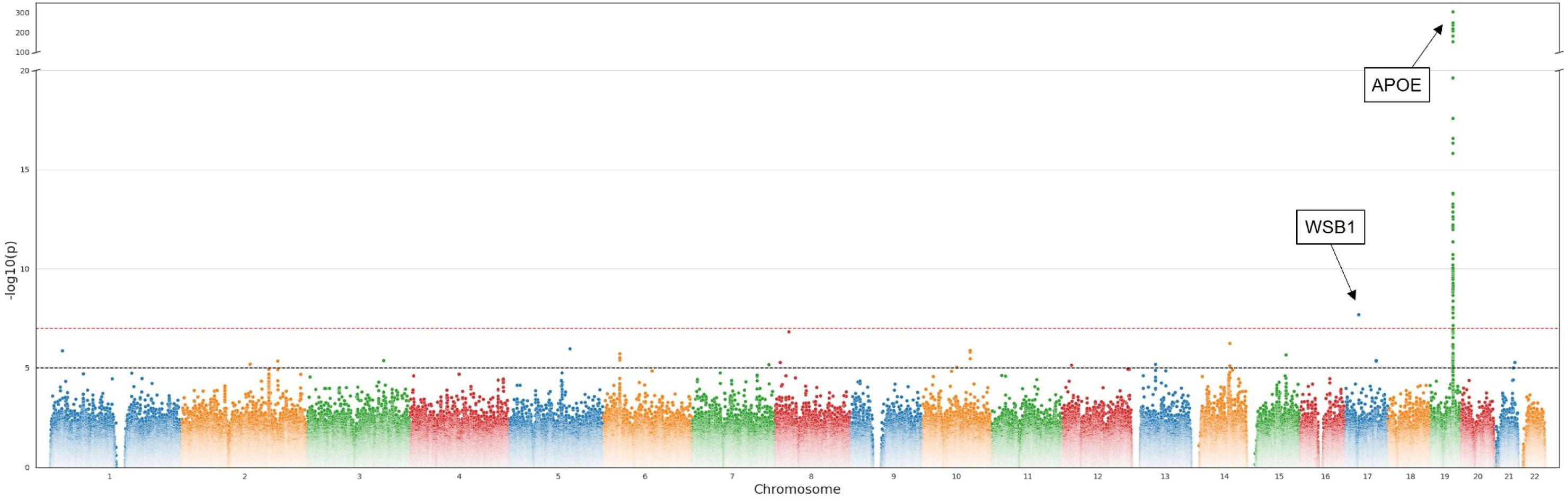
Manhattan plot showing results of GWAS using linear regression for AD and AD-by-proxy status and WGS-derived STRs on “Halldorsson-White-British” cohort. In the box, the nearest protein-coding gene according to GENCODE V47 is annotated. Horizontal red dashed line indicates the genome-wide significance threshold of 1.01E-07, whereas the black dashed line indicates the suggestive significance threshold of 1.00E-05. Note that y-axis is discontinuous and capped at 300. See supplementary Table 11 for more details on these results.

Next, we assessed why the novel genome-wide significant signal near *WSB1* did not also emerge in the GWAS using imputed STRs. Inspection of the imputed genotype data at this STR revealed that the allele giving rise to the association signal, did not pass QC in the imputed STR data (due to an excess of missing genotype data). LD mapping (±500kb) revealed no other SNPs or STRs in strong LD (r2>0.2) with the lead variant. Conditional analyses on this STR signal including the strongest SNP in the region (rs117222268 from ref. 3) revealed that the STR association remained genome-wide significant after adjustment (p=1.77E-08; Supplementary Table 12). While testing the associated *WSB1* STR in the “Halldorsson-Other White” subset did not show evidence for association with AD risk (Supplementary Table 10), we note that the number of AD cases in this portion of UKB data is exceedingly small (n=48 AD cases and n=887 AD-by-proxy). This combined with the fact that the frequency of the associated allele is relatively low (MAF=0.0145) substantially reduced the power of the replication analyses.

Of the 71 genome-wide significant WGS-derived STR signals located in the *APOE* region, we highlight STR chr19:44921083:A_11_:A_13_ which maps immediately adjacent to another STR (chr19:44921097-44921125:TTTA_n_) that was already reported to be associated with AD in previous work^19,27^. Conditional analyses using rs429358 led to a drop in the STR signal to p=0.01, suggesting that this STR does not elicit genetic effects over and above the well-known e4-allele. However, adjustment of the STR signals in this general region by *APOE* rs429358 led to a stark increase in effect size and association of the same STR for which a similar situation was observed in the imputed dataset (chr19:44910661, which may at least partially be due to the effect of the “e2-allele” in *APOE*, see above). Here, the p-value increased from 1.34E-01 before adjustment to 2.38E-08 after adjustment. These findings, again, suggest that the genetic architecture contributing to AD risk in the general *APOE* region may go beyond the effects of the e4-allele and deserves detailed fine mapping in future work.

Lastly, we investigated how the most promising signal near *ABCA7* performs in analyses using WGS-derived STRs. Since the lead imputed STR was not available among the WGS-derived STRs, we performed pairwise LD mapping in the ±500kb interval surrounding this STR and identified STR chr19:1037970:T_14_>T_11_ as a reasonable proxy (r2=0.65). In the WGS-GWAS, this variant showed nearly genome-wide suggestive evidence of association (p=7.68E-05, Beta=0.0114), supporting the overall notion that the AD association signal near *ABCA7* is largely STR-driven.

### Comparison of imputed vs. WGS-derived STRs

In an attempt to assess the general validity of imputed STRs we compared STR-based GWAS results to those using WGS-derived genotype data. As expected, the overall association evidence was less pronounced when compared to the discovery GWAS on imputed STRs (compare Figures 1 and 2), likely due to the much-reduced sample size of the WGS-derived vs. imputed dataset (n=95,201 vs. 295,551, respectively). Next, we repeated the GWAS analyses on imputed STRs in the WGS subset (n=95,201; Supplementary Figure 19) and assessed the correlation of effect sizes across both sets of analyses. We manually curated and compared association tests for all WGS-derived and imputed STRs with p<1.0E-05 in the respective GWAS (Methods). This resulted in n=84 matching variants where GWAS-derived effect size estimates showed a very strong correlation (Spearman’s r=0.99, p=8.77E-79; Supplementary Figure 20 and 21). These comparisons emphasize the overall high quality of the STR imputation process.

### Delineating potential functional mechanisms of AD-associated STRs

As stated above, most of the STRs identified to be associated with AD risk are located within or very close to ORFs of protein coding sequences. Hence, a potential functional consequence of the associations may be an effect on gene expression. As a first step to assess this hypothesis, we scrutinized all highlighted STRs regarding their potential to influence either DNA methylation (DNAm) or gene expression in *post-mortem* human brain samples (entorhinal cortex; n=145 and 173^28,29^).

To this end, we analyzed all CpGs within a ±1Mb window (i.e. those in *cis*) from genome-wide significant STR loci. After analysis, the resulting P-values were FDR-controlled using the Benjamini-Hochberg procedure (q=0.05)^30^. Using this threshold, 6 STR loci showed associations with 31 CpG sites in the brain (of which 26 were unique; Supplementary Table 13). One of the most interesting STR meQTL findings (summarized in Supplementary Table 13) was observed with CpGs on chromosome 7q22 located between *NYAP1*, a gene that is predicted to be involved in neuron projection morphogenesis^31^, and *PILRB*, a gene involved in immune system function (https://www.uniprot.org/uniprotkb/Q9UKJ0) that has already been linked to AD in several previous studies^32,33^. The meQTL association is driven by STR chr7:100326718:CT:C showing strong association with two CpGs (cg06214670 [p=4.63E-09] and cg03579757 [p=3.27E-09]). Furthermore, several meQTL associations were observed with CpGs mapping in or close to *HLA*-related genes (chr. 6p22) in particular *HLA-DRB1* (e.g. cg08845336 [p=1.33E-14] located in exon 2 of *HLA-DRB1*) which is involved in the adaptive immune response and represents an emerging candidate gene for AD and other neurodegenerative disorders^34–36^. Lastly, we also found strong evidence for an STR-based meQTL in a proximal enhancer element of the second intron of *SNX32* (cg15531562 [p=7.07E-13]), a gene involved in the regulation of neurite outgrowth^37^ representing one of the novel STR-related AD loci in our discovery GWAS analyses.

Next, we computed STR-based expression QTL (eQTL) analyses using gene expression profiles derived from RNA sequencing experiments conducted in the same brain samples (n=173; Aslam et al., in preparation; and reference 29). For each STR, we probed for associations with genes within a ±1Mb window, again using FDR adjustment^30^. In general, there was a good correspondence between meQTL and eQTL results and four STR-mRNA pairs attained study-wide significance after FDR correction (Supplementary Table 14). Of these, the strongest and most consistent eQTL signals were, again, observed with genes in the HLA region on chromosome 6p22. Future work in larger brain-based datasets needs to follow up on these meQTL and eQTL leads.

## Discussion

In this study, we conducted the first STR-based GWAS for AD in a large collection of ∼330,000 UKB individuals. In analyses using imputed STR data, we identified 14 loci showing evidence for genome-wide significant association with AD risk, the strongest effects elicited by STRs in the *APOE* region. Replication testing in a subset of ∼20K UKB individuals provided independent support for all but one of these 14 loci. Among all genome-wide significant signals in the UKB cohort, there was one where STRs (and not SNPs) represent the lead signal (*ABCA7*), and three where STRs show noteworthy contributions to the SNP-driven associations with AD risk (*HLA-DRB1, MINDY/ADAM10*, and *APOE*).

GWAS analyses using WGS-derived STRs led to the identification of one additional genome-wide significant locus (near *WSB1*) previously not highlighted by SNP-based GWAS. Post-GWAS analyses suggest that STRs make up at least 3% of the overall genetic heritability of AD in this dataset. Lastly, aligning our top STRs in genome-wide DNAm and transcriptome profiles from independent human brain samples suggested that several of the identified STRs may unfold their effects by impacting the expression of nearby genes.

Our study reports three main novel findings. First, we detected two new potential STR-based GWAS signals for AD, i.e. near *OVOL1/SNX32* on chr. 11q13, and near *WSB1* on chr. 17q11. None of the known AD SNP-based GWAS results ^3^ mapped within ±0.5Mb from these loci, qualifying them as novel signals. Despite the fact that both of these loci contain promising AD candidate genes (see below), we note that neither was independently replicated in the “Other White” subset of the UKB dataset (but there was support from other descent groups for *OVOL1/SNX32*; Supplementary Table 5). The first signal was implied by an imputed STR (chr11:65810443:TCA:T; p=7.01E-08) near *OVOL1* (∼13.2kb upstream; encoding) and *SNX32* (∼23.5kb downstream). While our literature search retrieved no obvious link to AD for *OVOL1*, the situation is different for *SNX32*. This gene encodes sorting nexin 32 which is very highly (and predominantly) expressed in brain (https://www.gtexportal.org/home/gene/SNX32 and https://www.proteinatlas.org/ENSG00000172803-SNX32). The STR associated with AD also represents one of the strongest brain-based meQTL signals of this study with CpGs annotated to *SNX32* (p=7.07E-13; Supplementary Table 13). Interestingly, this gene was recently highlighted as a novel AD candidate protein in a proteome-wide association study (PWAS) where it showed evidence pointing toward a causal involvement in AD pathogenesis^38^.

Functionally, SNX32 was shown to be involved in neuronal differentiation in neuroglial cells *in vitro*^37^. The second locus maps to chr. 17p22 and is elicited by a WGS-derived TG dinucleotide STR. It maps ∼29.4kb upstream from the *WSB1* gene into an H3K27Ac mark, indicating an active enhancer element (http://genome.ucsc.edu). *WSB1* mRNA is ubiquitously expressed, including brain (https://www.gtexportal.org/home/gene/WSB1). Functionally, the encoded protein is a member of the WD-protein subfamily and a probable substrate-recognition component of a SCF-like ECS (Elongin-Cullin-SOCS-box protein) E3 ubiquitin ligase complex which mediates the ubiquitination and subsequent proteasomal degradation of target proteins (www.uniprot.org/uniprotkb/Q9Y6I7). As such, it is involved in the proper functioning of the ubiquitin-proteasome system (UPS), which also plays a central role in the accumulation of pathogenic proteins in AD, including Aβ and tau^39^. Due to its low MAF (∼0.01) and the complex genomic architecture in this chromosomal, independent replication testing was either not informative or not possible.

The second major novel finding of our study highlights two previously established AD genetic loci, i.e. near *ABCA7* (chr. 19p13) and *MINDY2/ADAM10* (chr. 15q21), for which STRs either represent the lead association signal or make strong contributions to known SNP effects. First, SNPs in *ABCA7* have long been established to show an association with AD risk^40^. Various comparative post-GWAS analyses suggest that STRs – perhaps more so than SNPs – may be the drivers behind this signal. A similar observation was made for the second locus, i.e. the region harbouring *MINDY2* and *ADAM10*. Although the genetic evidence points more towards *MINDY2*, the second gene in this region, *ADAM10* (encoding α-secretases a disintegrin and metalloprotease 10), which functions as constitutive α-secretase cleaving amyloid-β protein precursor (APP) protein in the non-amyloidogenic pathway^41,42^, appears as the more compelling candidate.

The third main finding is related to various STR-based association signals within the *HLA/MHC* region. Here, the GWAS results on both the imputed and WGS-derived STRs suggest association signals that remain stronger after conditioning on top GWAS SNPs than vice versa, suggesting that STRs potentially make a more pronounced contribution to AD risk than SNPs. In addition, the same STRs also show strong and consistent associations with both DNAm and gene expression in human entorhinal cortex samples. All of these association signals are led by STR chr6:32611487:TCTTTCTTTC:T, which decreases risk for AD (beta=-0.0090, p=1.23E-08) and DNAm (at cg26036029; beta=-0.1391, p=3.57E-12) while increasing the expression of *HLA-DQA2* (transcript ENST00000374940; beta=0.2810, p=2.07E-07) in brain. In addition, strong eQTL signals are also observed for two WGS-derived STRs in the region (p=1.07E-06). Thus, there is remarkable consistency of findings in this region on the genetic (GWAS), methylomic (meQTL), and transcriptomic (eQTL) levels, strongly supporting the genuineness of this finding.

Despite its strengths, our study also has several potential limitations. First, our main and most powerful GWAS analyses (in ∼330,000 individuals) are based on imputed STR genotypes.

While comparisons between AD-associated STRs from imputation and WGS indicated a high correlation of GWAS test statistics (confirming the validity of our main findings) it is important to note that current STR imputation paradigms do not yet achieve the same high quality achieved for imputation of SNPs^16^. Second, to streamline the statistical analyses, we split multi-allelic STRs into bi-allelic STRs which might have reduced our power to detect association signals at some loci. We note, however, that this procedure does not invalidate any of the bi-allelic association signals reported herein. Third, the proxy phenotyping approach used was recently shown to be the source of some bias in the context of GWAS, mostly by potentially inflating or skewing estimates of genetic correlations between traits^43^. It is important to emphasize that only a negligible bias was detected for the *discovery* of GWAS loci^43^, which is how proxy phenotyping was used here. Fourth, our analyses mainly focus on UKB individuals of European (“white”) descent. By a margin, these decent groups represent the largest subsets within the UKB release included here. While several of our top STR GWAS signals either directly or indirectly replicate in other ancestries available in UKB, these analyses are based on comparatively small sample sizes (containing maximally ∼400 AD and AD-by-proxy cases). Lastly, while we were able to delineate some initial hypotheses on putative mechanisms underlying our top STR-based association signals by using genome-wide DNAm and transcriptome profiles in an independent set of human brain samples, this latter dataset is comparatively small (n_max_∼173) and only includes one brain region (entorhinal cortex).

In conclusion, our study constitutes the first large-scale STR-based GWAS for AD risk in the field. In addition to detecting two potentially novel and functionally interesting AD loci (near *SNX32* and *WSB1*), our results suggest that STRs may strongly contribute to the genetic effects at four additional loci (*ABCA7*, *MINDY2*/*ADAM10*, *HLA-DRB1,* and *APOE*).

Furthermore, we estimate that STRs account for at least 3% of the genetic variance underlying AD. Future studies, ideally using directly genotyped STRs in large AD datasets, are needed to confirm our results and to further delineate the likely sizeable role that STRs play in the genetic makeup of AD.

## Methods

### Human subjects

Collection of the UKB data was approved by the Research Ethics Committee of the UKB obtained under application ID 81874. All UKB participants provided informed consent. The UKB contains data for 502,364 samples, of which 488,377 have array SNP data available (Field ID-22418). We checked the 488,377 samples for mismatches in reported and genotyped sex, aneuploidy, and heterozygosity outliers using genomic QC-tests that are provided within the UKB (Field IDs 22001, 22019, and 22027), removing 1,805 samples in total. Of the remaining 486,539 samples, 3,783 are classified as AD cases based on ICD-10, while 482,539 are non-cases. Samples without an ICD-10 diagnosis of AD were further subjected to “proxy phenotyping” (see below). To this end, we check whether they provided information about parental age and AD status. This information was not available for 93,260 samples which were subsequently removed from all analyses. Next, we checked all remaining samples for possible relatedness using UKB data field 22021. Participants with no relatives identified were kept (n=273,321) and samples with 10 or more relatives were removed (n=169) without further analyses. For the remaining 119,572 samples with at least one but less than ten relatives according to UKB data field 22021, we ran kinship removal tests based on KING algorithm provided by PLINK2 (v2..00a3LM, www.cog-genomics.org/plink/2.0/)^44^, prioritizing cases and proxy-cases above controls, removing 59,447 additional samples. This procedure resulted in a final dataset of 333,446 samples after QC.

The 333,446 samples were split into groups by self-reported ethnic background (UKB field ID 21000_i0). Participants labeled as “White” are divided into “White-British” (n=295,551; “discovery” sample in GWAS) and “Other-White” (n=20,840; “replication” sample) cohorts. Due to their sample size, other groups (“Asian”, “Black”, “Chinese”, “Mixed”, “No_Answer”, and ‘Other’) were not further subdivided. All samples were additionally filtered according to the availability of WGS-derived STR genotypes as determined by Halldorson et al.^26^, leaving 107,289 samples for that analysis arm.

A visual summary of this workflow can be found in Supplementary Figure 1.

### Procedure for “proxy phenotyping”

UKB provides information about participants’ health records in form of ICD-10 codes. Samples with any form of AD diagnosis by ICD-10 (Code F00: ‘Dementia in Alzheimer’s disease’; Code G30:’ Alzheimer’s Disease’) were considered to be “diagnosed AD cases” (n=3,783).

Proxy-Scores were then assigned to the remainder of the dataset based on self-reported parental AD-status. Each participant not categorized as “diagnosed AD case” was assigned an “AD proxy score” ranging from 0-2. The algorithm to calculate proxy scores was derived from Jansen et al.^45^. Proxy scores are composed of parental risk-scores for AD, ranging from 0-1 per parent, where parents that have a recorded form of any AD are scored as 1. Otherwise, they get a risk score of their age relative to 100, capped at 0.32 to not exceed the maximum prevalence of AD as reported in Herbert et al.^46^, such that *risk _parent_* = *min*((100-*age_parent_*)/100, 0.32) and *proxy_score= risk_mother_ + risk_father_*. Next, scores <u>≤</u> 0.64 (i.e. samples without any parent affected) were considered as “controls” while scores between 0.64 and 2 (i.e. samples with at least one affected parent) were considered as “proxy cases”.

To account for the certainty of the disease status, “diagnosed AD cases” were assigned a proxy score of 4, i.e. they carry twice the weight as the highest-scoring proxy AD cases in the subsequent association analyses. Note that this differential weighting is not implemented in the procedure by Jansen et al., who assign equal weights to proxy cases with two affected parents and cases with an ICD-10 diagnosis of AD.

### STR data processing and quality control

<u>STR imputation and QC:</u> For STR imputation, we followed the approach published by^11,15^. This is based on genome-wide SNP genotyping data released by UKB (field ID=22418) and the 1kGP SNP-STR haplotype panel released by Ziaei Jam et al.^16^. Before imputation, SNPs are filtered for genotyping efficiency (exclude all with <98%) and for minor allele frequency (exclude all with <1%), followed by conversion to vcf-format and lifting from hg19 to hg38 using GATK LiftoverVcf^47^. To reduce computation time, samples were divided into batches of n=5000 before imputation and matched with the imputation panel using the conform-gt tool. STR genotype imputations were computed using Beagle V5.4^48^. After imputation, all SNP genotypes were removed, followed by splitting of multiallelic STRs into a biallelic format and filtering for imputation quality (exclude all variants with DR^2^ <0.7). Due to imperfections in the imputation catalog, the QC’ed data still contained genotypes on a number of non-STR variants, e.g. indels and SNPs, and duplicate STR entries, requiring manual inspection and removal. To facilitate this process, non-STRs were removed after GWAS from the set of either genome-wide significant or suggestive association signals.

<u>QC of WGS-derived STRs:</u> UKB also provides WGS-derived (i.e. non-imputed) STR data for 150,000 participants published in an UKB-analysis of WGS data by Halldorson et al. (2022) (Field ID 23365)^26^. Before analysis, the WGS-derived STR data were QC’ed and split into biallelic variants, as outlined above for imputed variants.

<u>QC of genome-wide SNP genotyping data:</u> For SNP-based GWAS analyses, imputed SNPs provided by UKB (Field ID 22828) were used. First, they were lifted from hg19 to hg38 using GATK LiftoverVcf^47^ and then filtered according to the following criteria using PLINK (v2.00a3LM, www.cog-genomics.org/plink/2.0/)^44^: All samples are split into the respective cohorts, variants with minor allele frequency below 1% and genotyping efficiency below 2% are filtered (--maf 0.01 --geno 0.02). Also, variants with deviations from HWE at P<5E-06 (--hwe 5e-06) in controls were removed.

### Genome-wide association analyses

Association tests were run for imputed STRs, imputed SNPs, and WGS-derived STRs in PLINK (v2.00a3LM) using linear regression models on the proxy phenotyping score (see above) adjusting for genetic ancestry (using the first 15 principal components [PCs] from a principal component analysis on LD-pruned non-imputed SNP genotype data) and sex. For the SNP-based data (i.e. imputed STRs and SNPs) we additionally adjusted for genotyping-array (field ID 22000). For the WGS-derived STR data, we adjusted for sequencing provider (200K-release; field ID 23080). To define a genome-wide significance threshold we estimated the number of independent (r^2^ <0.2 or more than 1500kb apart) imputed STRs to be n=334,605 and used this number for determining a Bonferroni-corrected significant threshold of α = 1.49E-07 (α = 0.05/334605). Similarly, for the WGS-derived STRs, we identified n=495,465 independent STR-variants resulting in a Bonferroni-corrected significant threshold at α = 1.01E-07 (α = 0.05/495,465).

After GWAS for imputed STRs, since the STR reference panel used for imputation^16^ contains several imperfections, we performed a manual check for all genome-wide significant and suggestive signals based on each STR’s genome coordinate. This led to the removal of 223 variants that were duplicated (with different IDs) and 370 non-STR variants (indels or SNPs), leaving 482 unique STRs (254 genome-wide significant STRs at 14 loci and 228 suggestive at 21 loci) (Figure 1, Supplementary Table 2).

### Replication analysis

To assess the validity of the primary GWAS findings in the discovery dataset of n=295,551 “White British” UKB individuals, we used the n=20,840 “Other-White” subset as replication cohort. These were analyzed using the same GWAS analysis paradigms as for the discovery computations. To count as “direct replication”, variants needed to show at least nominal evidence of association (p<0.05) and the same direction of effect as in the primary GWAS. Lastly, results from both discovery and replication datasets were combined by fixed-effect meta-analyses using PLINK (v1.90b7). In this context, we considered variants showing stronger statistical support (=smaller p-values) in the meta-analysis as compared to the discovery results but not fulfilling the above replication criteria as “indirect replications”.

### Fine mapping and conditional analyses

A range of analyses aimed at discerning SNP from STR effects were performed. This entailed running a SNP-based GWAS on the discovery dataset using imputed genotypes released by the UKB.

To identify lead variants within the imputed STR data, we used GCTA-COJO^49^. The same tool was used on combined SNP and STR GWAS summary statistics to distinguish STR and SNP signals in our results.

To identify lead STR variants, we determined the most significant variant within any given 250kb window by checking whether there was any variant with a smaller p-value in this vicinity.

In addition, we performed two types of conditional analyses to compare SNP vs. STR data: i) we adjusted the STR signal for the effects of the respective lead SNP (±500kb) per STR locus derived from the SNP-based AD GWAS by Bellenguez et al.^3^. This was done for all significant loci except *APOE*, which was excluded in the study by Bellenguez et al. Here, we adjusted for the well-established epsilon-4 AD risk allele (i.e. T-allele at rs429358). ii) We performed reciprocal analyses where the SNP association statistics in any of the top STR loci were conditioned on STR genotype. In case the top regional Bellenguez SNPs did not fulfill our variant QC-criteria (MAF <0.01 and genotyping efficiency <0.98) in the UKB SNP dataset, we searched for the next strongest SNP until the criteria were fulfilled.

### Heritability Analysis

Heritability analyses were run using the GCTA-LDMS method^50^, following the protocol provided by the authors. Here, a segment-based LD score is calculated first, based on which all variants are stratified into four LD groups. One advantage of this method is that it prevents LD bias among variants imputed from SNPs, which may occur using the regular GCTA algorithm^50^. To increase the precision of the resulting estimate and to improve computational efficiency, the heritability calculations were run on a subset of individuals within the discovery cohort of “White British” individuals, i.e. all n=2947 diagnosed AD cases and 15,000 randomly selected controls. Heritability analyses were run for SNPs and STRs individually as well as the combined SNP and STR data. Contribution of STRs to the total AD heritability was calculated as follows: (h(SNP+STR)-h(SNP))/h(SNP+STR).

### Comparison of GWAS results for imputed vs. WGS-derived STR variants

A direct comparison of imputed vs. WGS-derived genotypes is aggravated and often impossible due to the fact that the use of different toolkits for STR-imputation and STR-genotyping leads to discrepancies in variant-IDs, as well as naming of reference and alternative alleles. Therefore, we compared the effect sizes from GWAS analyses on the same subset of UKB individuals (n=95,201; “Halldorsson subset”). All STRs passing suggestive significance in either imputation-or WGS-based GWAS were matched by position (n∼200) and manually curated with respect to “true” allelic matches. The resulting, uniquely matching variants were then subjected to Spearman’s rank-based correlation analysis.

### Methylation quantitative trait locus (meQTL) analysis

Targeted meQTL analyses were performed using genome-wide DNA methylation (DNAm) profiles assayed using the “Infinium MethylationEPIC” array (Illumina, Inc.) on DNA extracts from human brain tissue (entorhinal cortex, Brodmann area BA28) of 145 AD cases and controls from the Oxford Project to Investigate Memory and Aging (OPTIMA) dataset^28^. STR genotypes were imputed from genome-wide SNP genotyping data (Global screening array [GSA]; Illumina, Inc) using the same procedure outlined above for the UKB samples. Details on the OPTIMA dataset, as well as the procedures used for DNAm profiling can be found in Sommerer et al.^28^. For this study, only CpGs within a ±1Mb window from genome-wide significant STR loci were considered. Details regarding the computational packages used, QC, and the workflow of the general meQTL analyses can be found in Ohlei et al.^12^.

Briefly, we used the R-package “MatrixeQTL” applying linear regression models adjusting for sex, the first two principal components (PCs) of the genotype data, and the first three PCs of the DNAm data (m.s. in preparation).

### Expression quantitative trait locus (eQTL) analysis

Targeted eQTL analyses were performed using transcriptome-wide gene expression data derived from total RNA sequencing in the same dataset of OPTIMA brain samples (n=173) also used for the meQTL analyses (for details on processing and QC of these RNA sequencing data see reference 29). Analytical procedures for the eQTL analyses were equivalent to those described for the meQTL analyses above, i.e. we applied linear regression models including sex, the first four principal components of a PCA assessing genetic ancestry, and the first two principal components of RNA levels as covariate.

## Data availability

Access to the UK Biobank resource is available via application (https://www.ukbiobank.ac.uk/). Here all data will be uploaded within 6 months after publication (Application Number 81874) and will be available to any individuals with an approved UKB project. Summary statistics will be made public upon publication and on reasonable request.

## Code availability

No new software was developed for the analyses of this project. Instead, all software tools were previously published. Relevant references as well as parameters for their execution are provided in the methods section.

## Acknowledgments

This research has been conducted using the UK Biobank Resource under Application Number 81874. We acknowledge the Oxford Brain Bank, supported by Brains for Dementia Research (BDR) (Alzheimer Society and Alzheimer Research UK) and the NIHR Oxford Biomedical Research Centre. In addition, this work was funded by the Deutsche Forschungsgemeinschaft (“STaR-AD” project to V.D. [2354/1-1] and L.B. [BE2287/9-1) and the Cure Alzheimer’s Fund as part of the Alzheimers Genome Project^TM^ (to V.D. and L.B.). The authors thank Dr. M. Volkan Cakir and Ms. Ida Happel who supported some of the analyses performed in this study, and Prof. Bernhard Haubold for helpful discussions.

## Author contributions

Overall study design: V.D. and L.B. Data acquisition & processing: D.G., D.P., K.M., L.P., C.M.L., R.E.T., V.D., L.B. Statistical analyses: D.G., O.O., M.M.A., V.D. Interpretation of findings: D.G., R.E.T., V.D., L.B. Writing of first draft: D.G., V.D., L.B. Review, editing, and approval of final draft: all authors.

## Supplementary information

Supplementary Figures 1-21 and Supplementary Tables 1-14.

## Materials & Correspondence

Prof. Lars Bertram

