## Supplementary Figures for "GWAS on short tandem repeats identifies novel genetic mechanisms in Alzheimer’s disease"

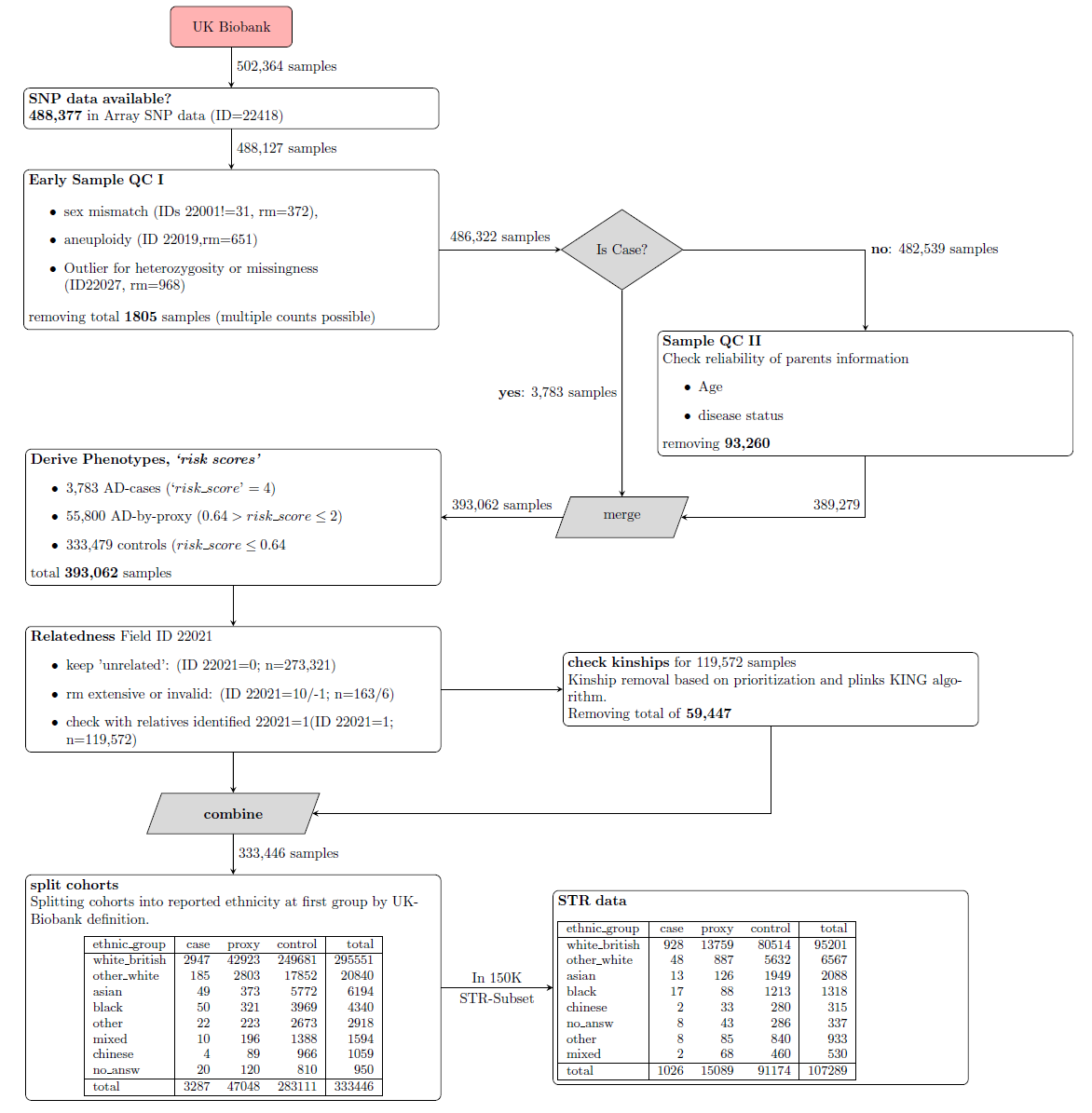

**Supplementary Figure 1**. Flowchart of data extraction and filtering implemented to identify eligible sample sets.

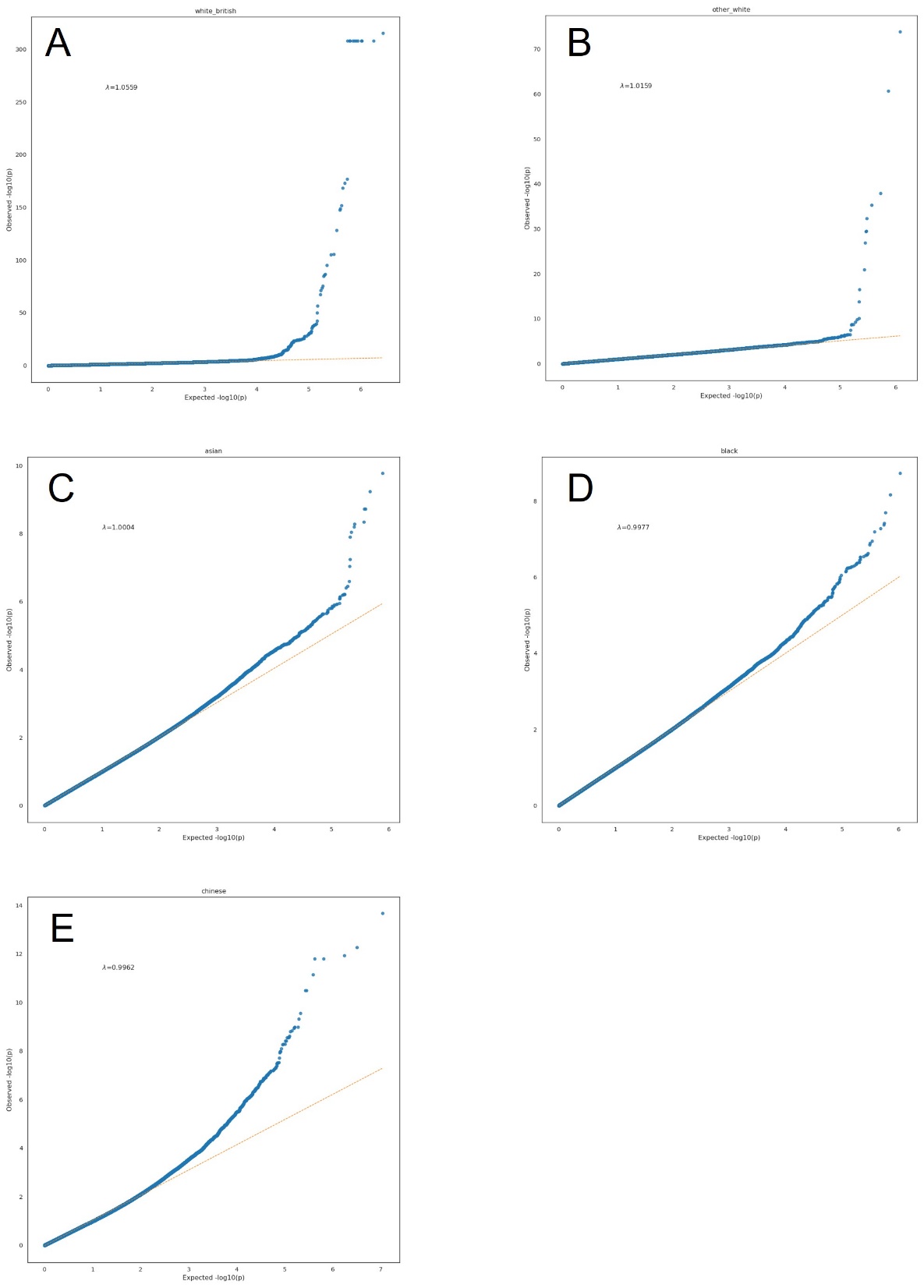

**Supplementary Figure 2**. QQ plots of imputed STR association results per cohort. The expected –log10 transformed p-values are displayed on the x-axis, while the observed –log10 transformed p-values are displayed on the y-axis. A - “White-British”, λ=1.0559; B - “other-White”, λ=1.0159; C - “Asian”, λ=1.0004; D - “Black”, λ= 0.9977; E - “Chinese”, λ= 0.9962.

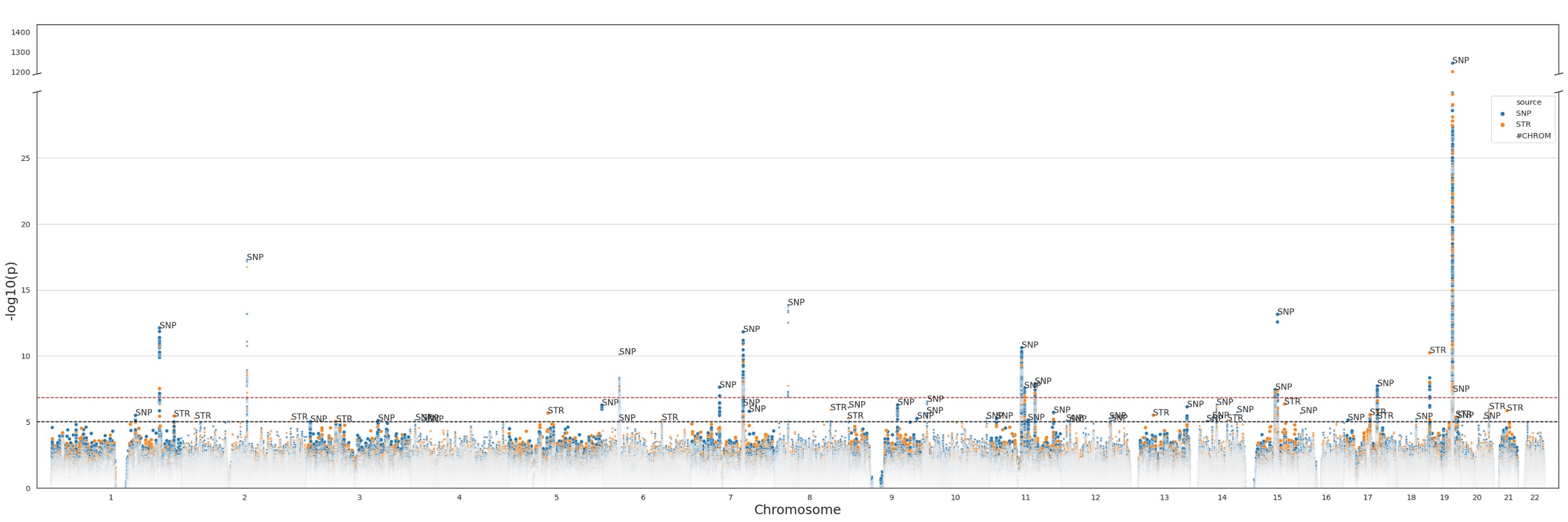

**Supplementary Figure 3.** Overlay of SNP-based (blue) and STR-based (orange) GWAS results for the White-British cohort. “SNP” and “STR” labels show the respective lead variant (by p-value) in peaks showing genome-wide significant and suggestive evidence for association.

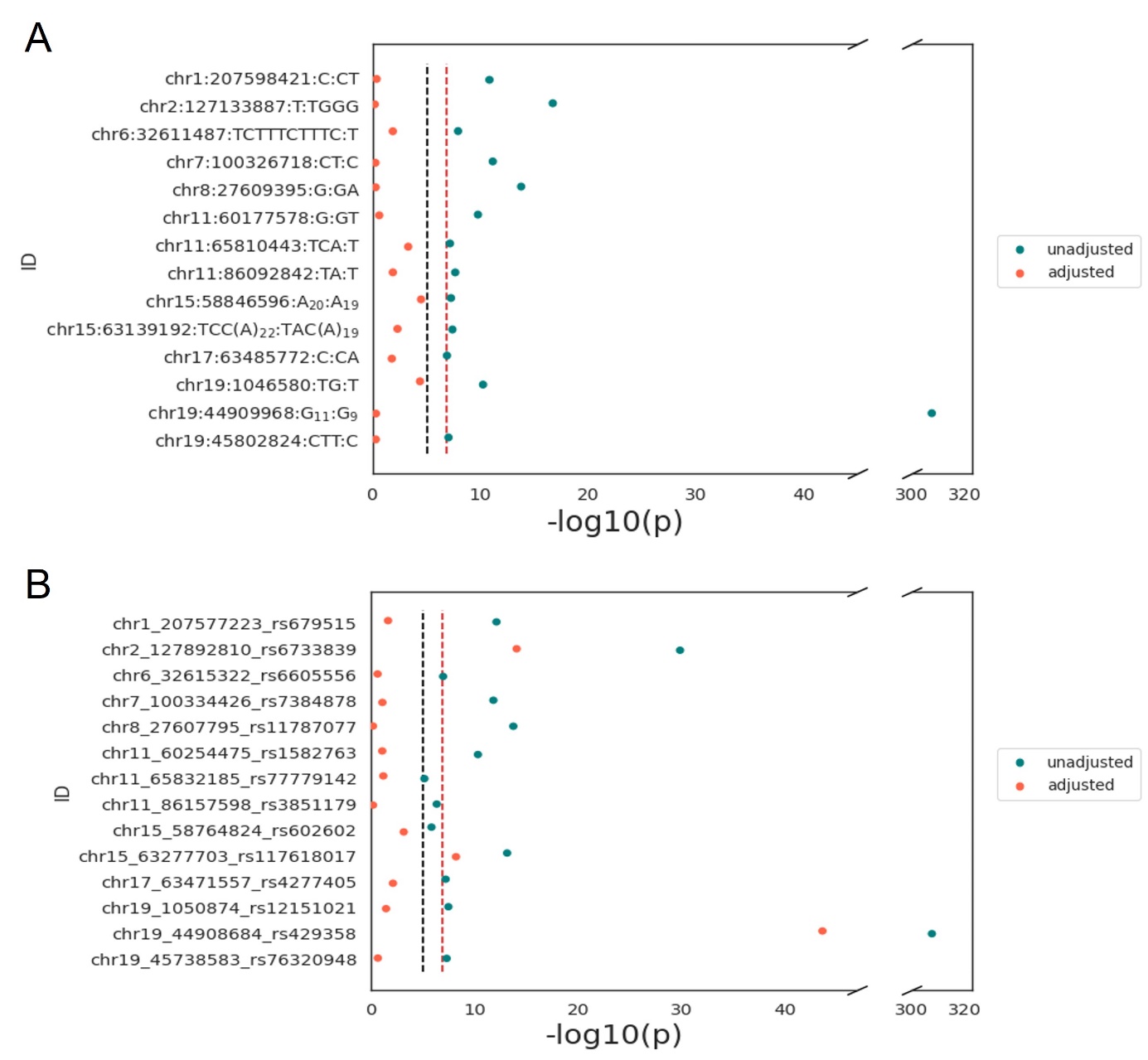

**Supplementary Figure 4.** Summary results of conditional analysis of (**A**) genome-wide significant imputed STRs (top hit per locus) with their corresponding SNP and (**B**) reciprocal conditioning of SNP with STR (Methods).

SNPs used for the conditioning are the lead SNPs at the corresponding locus (within ±500kb) from the largest AD GWAS published to date (Bellenguez et al., 2022). The exception is chr11:65810443, where the SNP with the lowest p-value was used, since there were no genome-wide significant SNPs in the given region. For the *APOE* region, which was excluded in the study by Bellenguez et al., we adjusted the STR-signal at chr19:44909968 for the well-established *APOE* e4-allele (i.e. T-allele at rs429358). For the STR-signal at chr19:45802824 (which was also excluded in Bellenguez et al.) we adjusted for rs76320948, which was identified as lead variant by Jansen et al., 2019. The vertical dashed lines indicate the genome-wide significance threshold of (p<1.49E-07; red) and the suggestive threshold (p<1.00E-05; black). Blue dots: -log10(p) values before adjustment, orange dots: -log10(p) values after adjustment.

**Supplementary Figures 5 - 18.** Results of conditional analysis of genome-wide significant imputed STR locus with corresponding lead SNP.

The lead STR-signals were adjusted for the corresponding SNPs at the given locus (left panel). SNPs utilized for adjustment were the lead SNPs from the largest AD GWAS published to date (Bellenguez et al., 2022), or, if unavailable, the SNPs showing the lowest p-value within ±500kb also passing early QC steps performed to prefilter the UKB datasets (i.e. MAF < 0.01 and missing genotyping call rate above 0.02). For the *APOE* region which was excluded in Bellenguez et al, we adjusted for the well-established AD risk allele at rs429358 and SNP rs76320948 from Jansen et al., 2019 for the region around the STR-signal at chr19:45802824. Right panels show the results of reciprocal conditioning of SNP-signals with relevant STR genotypes.

The horizontal dashed lines indicate the genome-wide significance threshold (p<1.49E-07; red) and the suggestive significance threshold (p<1.00E-05; black). Orange dots – the lead STR/SNP before conditioning. Orange crosses – the lead STR/SNP after conditioning.

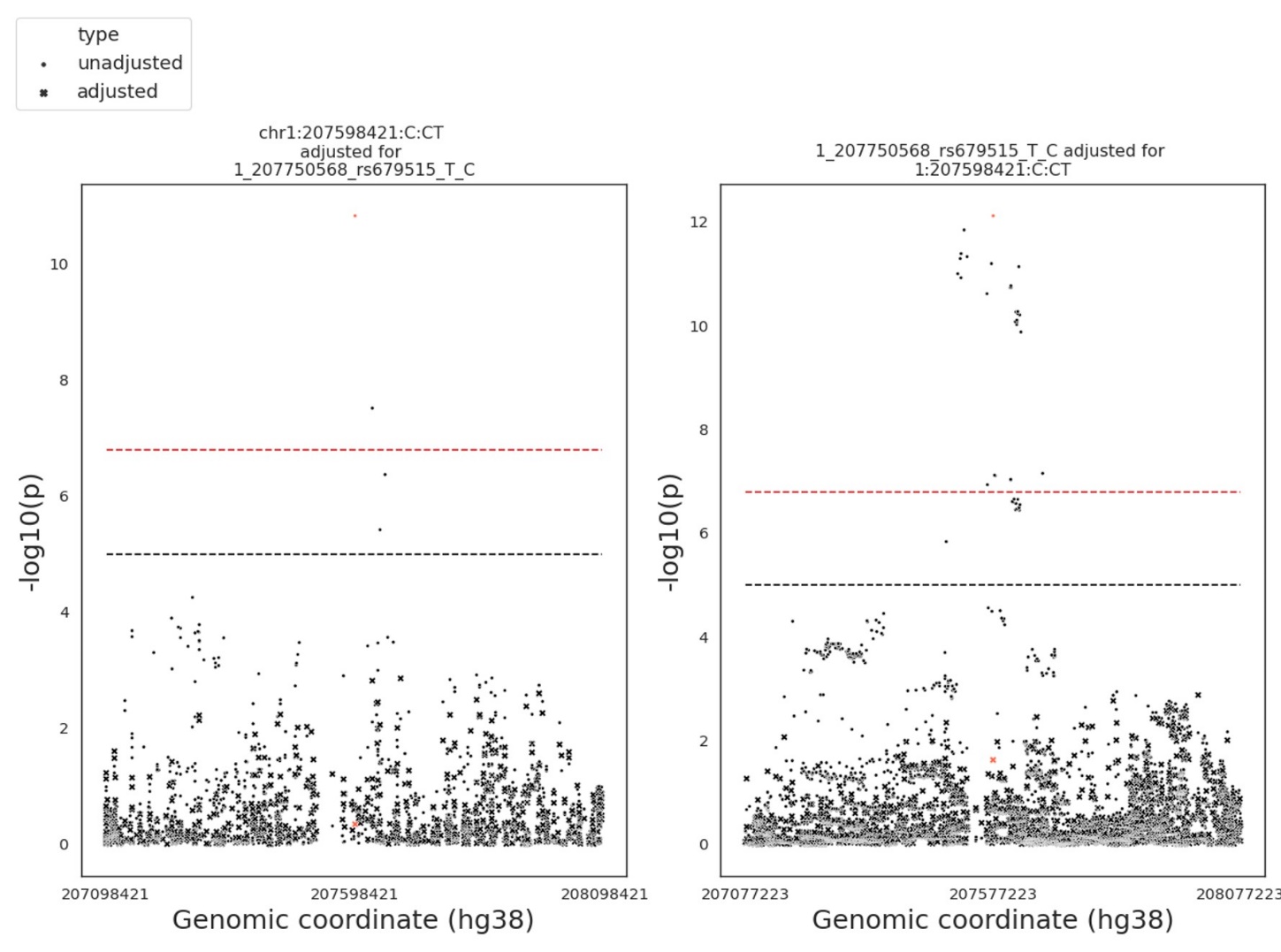

**Supplementary Figure 5.** STR-SNP pair: chr1:207598421:C:CT - rs679515.

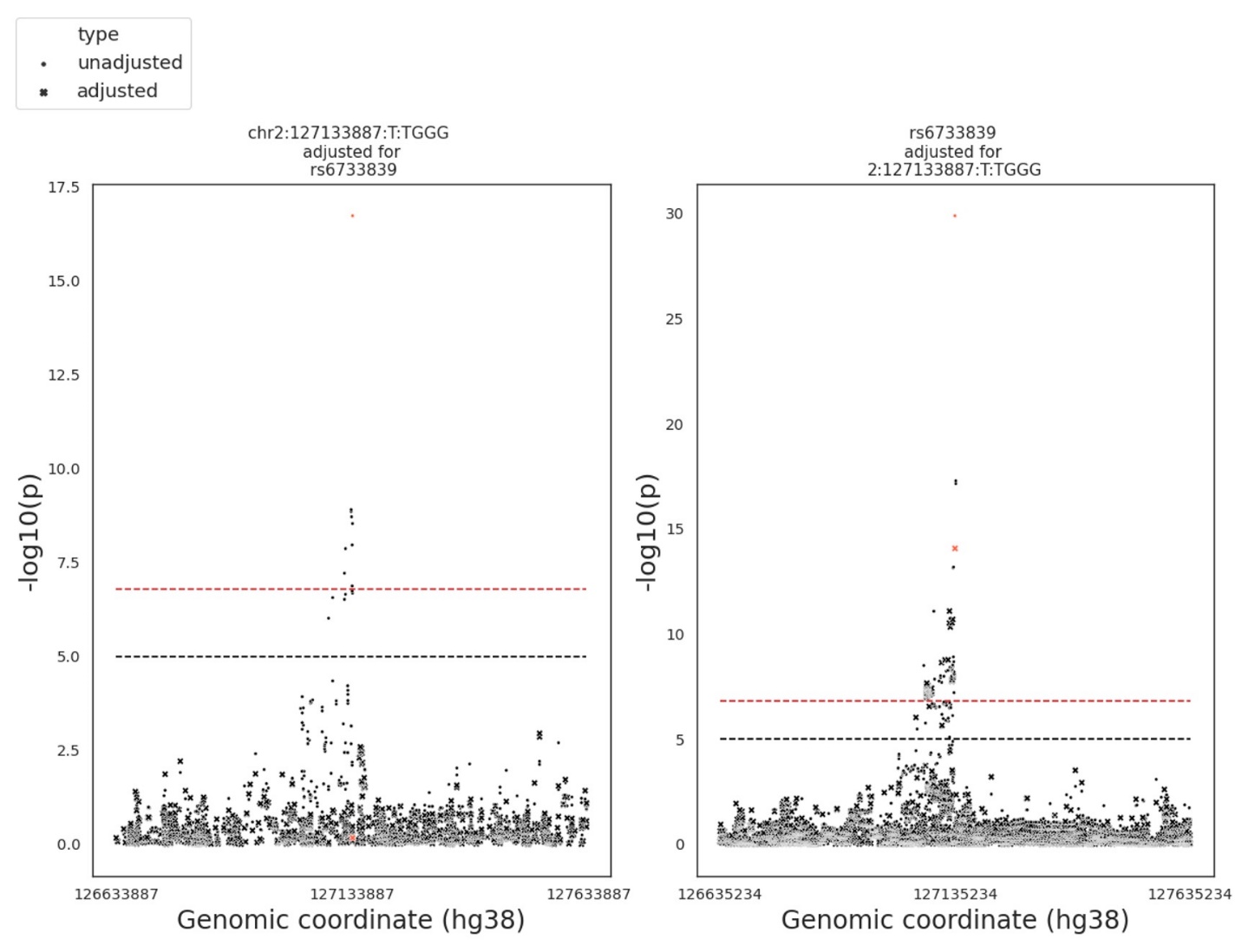

**Supplementary Figure 6.** STR-SNP pair: chr2:127133887:T:TGGG - rs6733839.

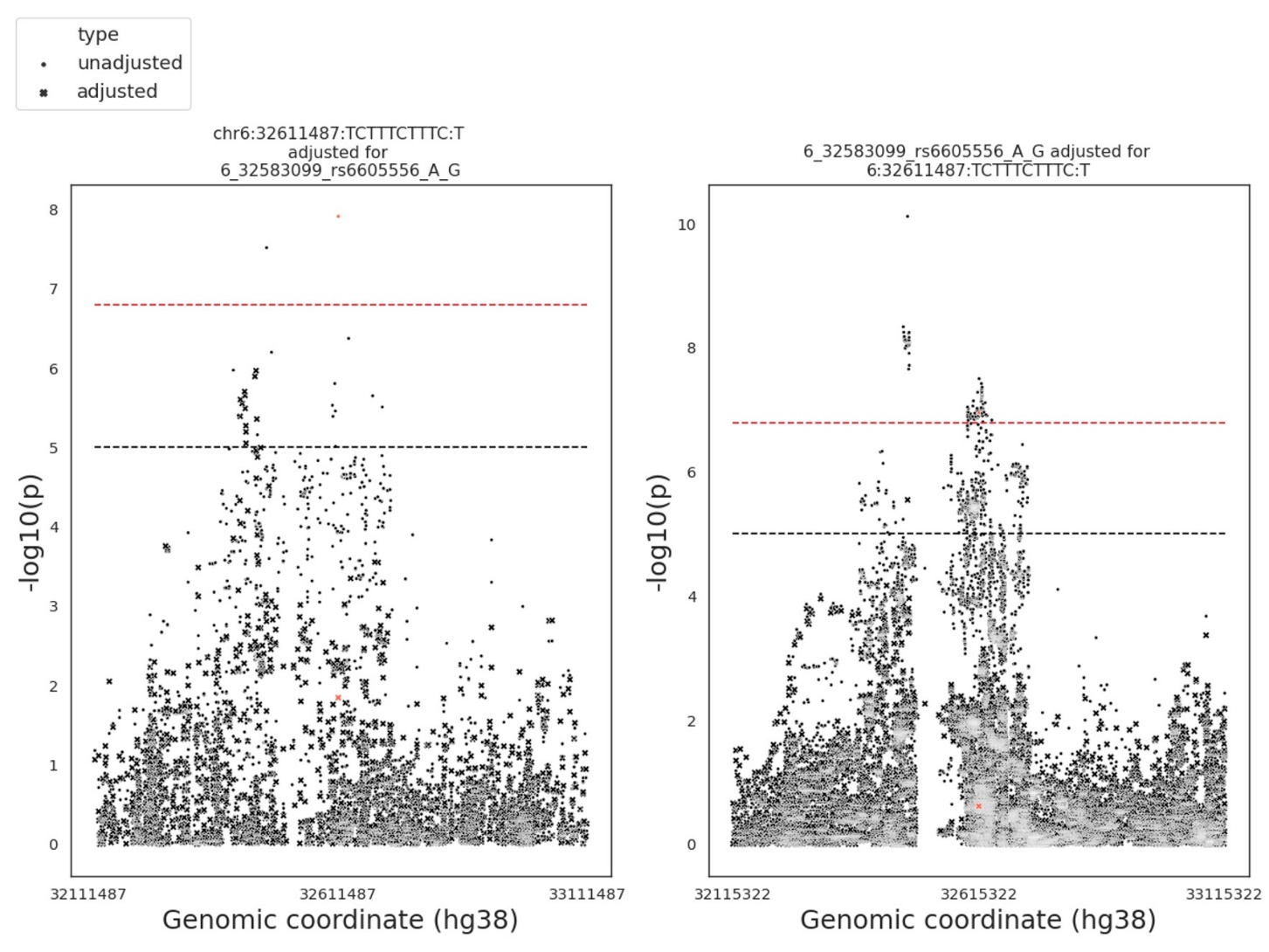

**Supplementary Figure 7.** STR-SNP pair: chr6:32611487:TCTTTCTTTC:T - rs6605556.

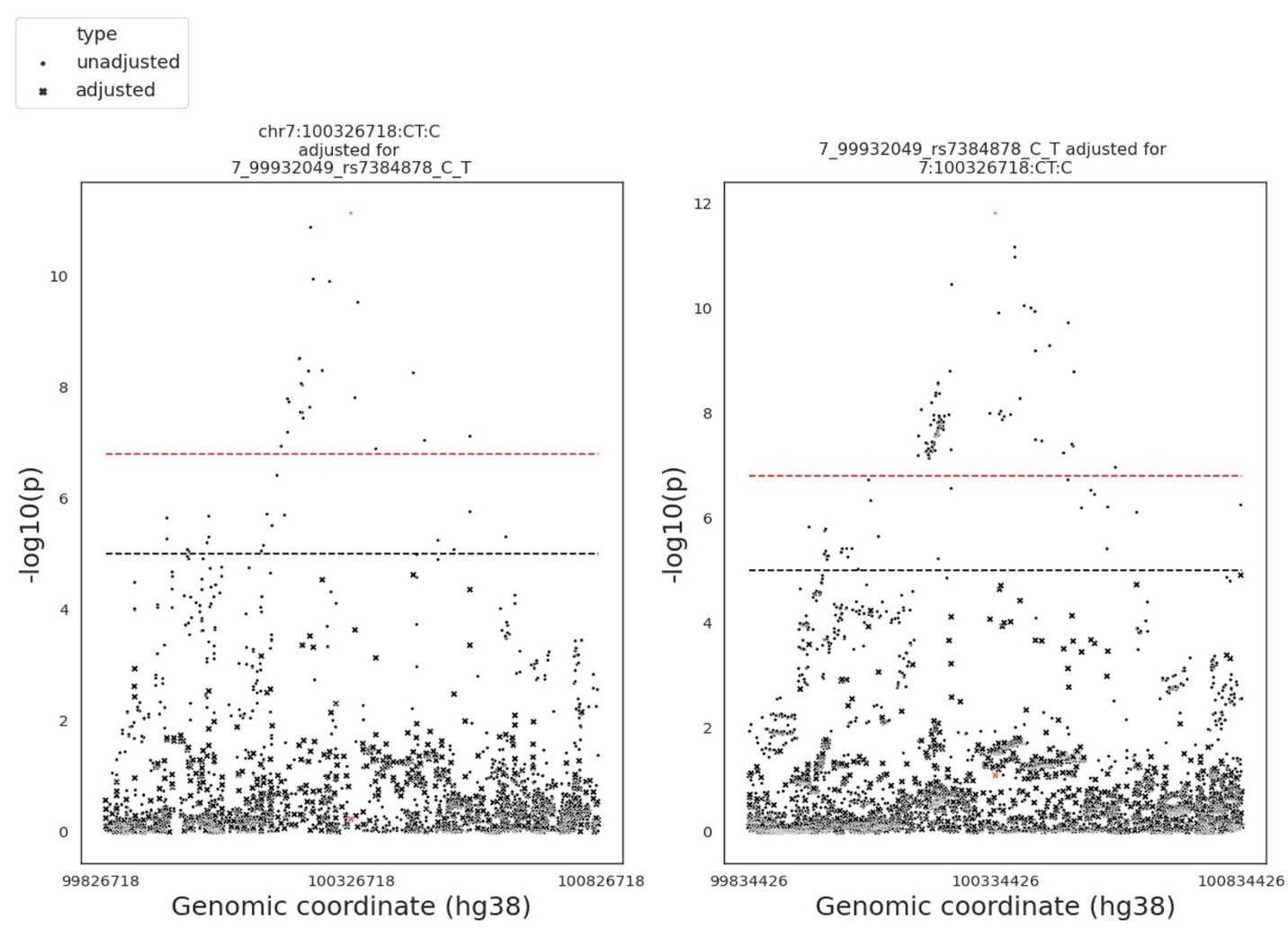

**Supplementary Figure 8.** STR-SNP pair: chr7:100326718:CT:C - rs7384878.

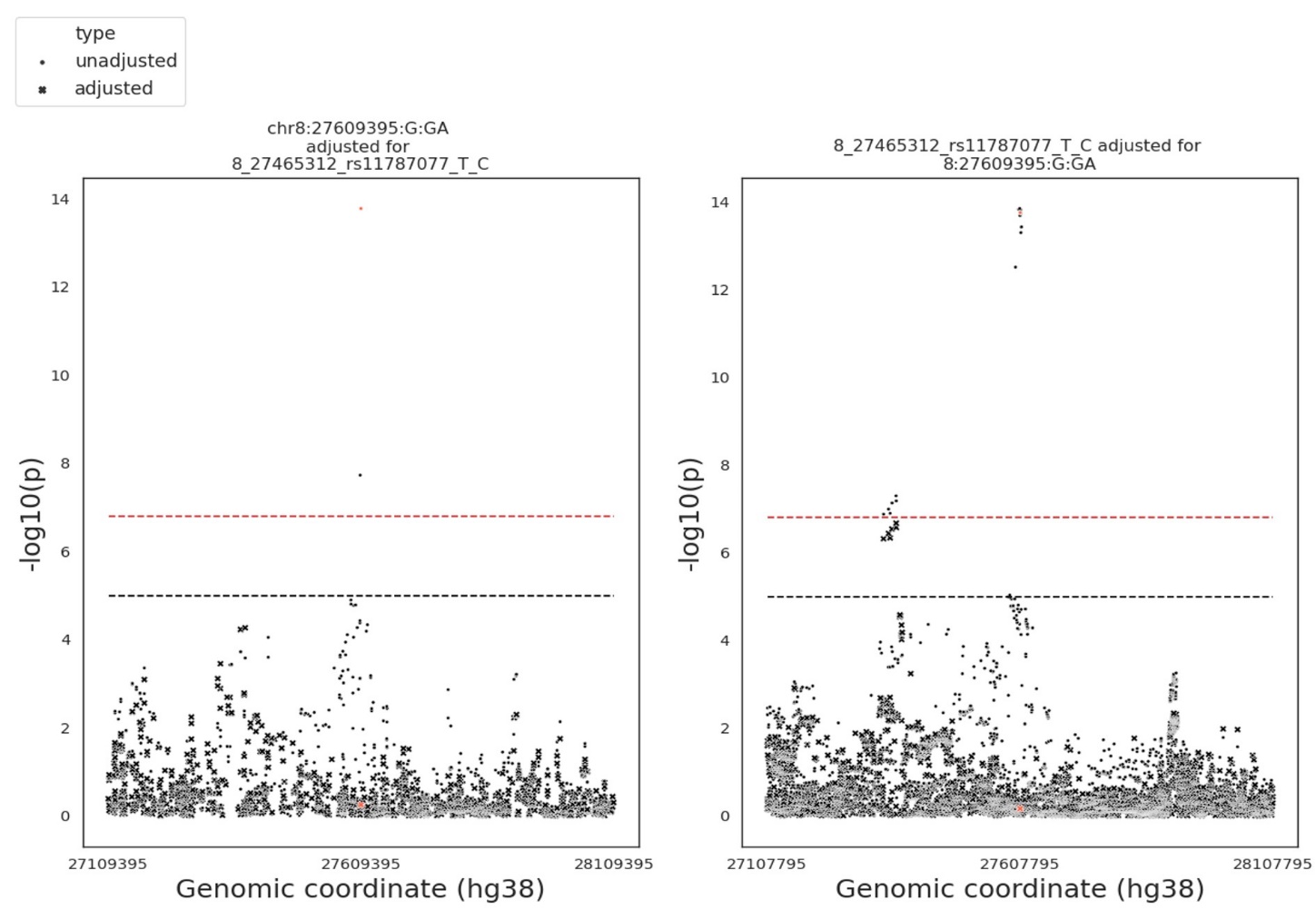

**Supplementary Figure 9.** STR-SNP pair: chr8:27609395:G:GA - rs11787077.

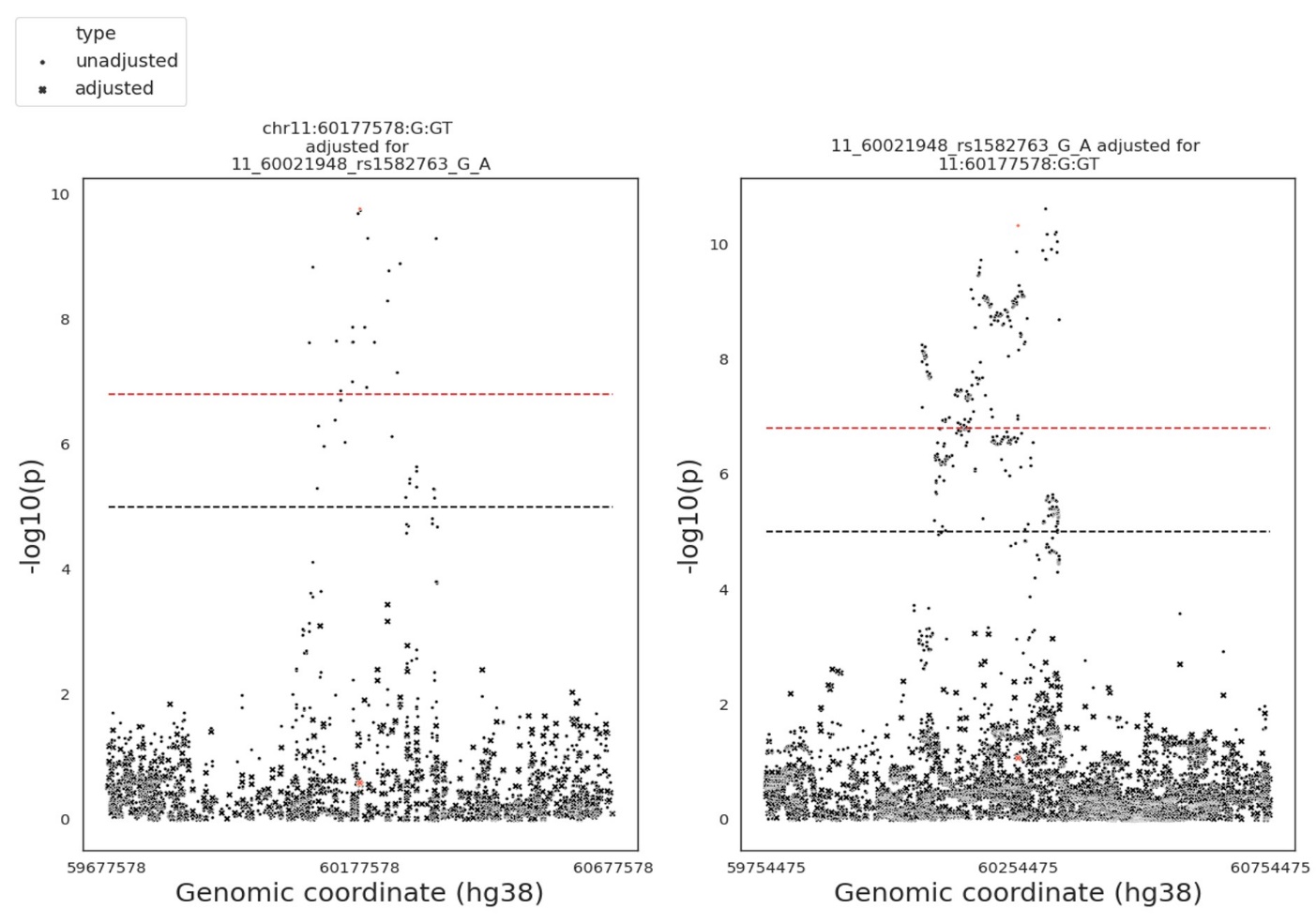

**Supplementary Figure 10.** STR-SNP pair: chr11:60177578:G:GT - rs1582763.

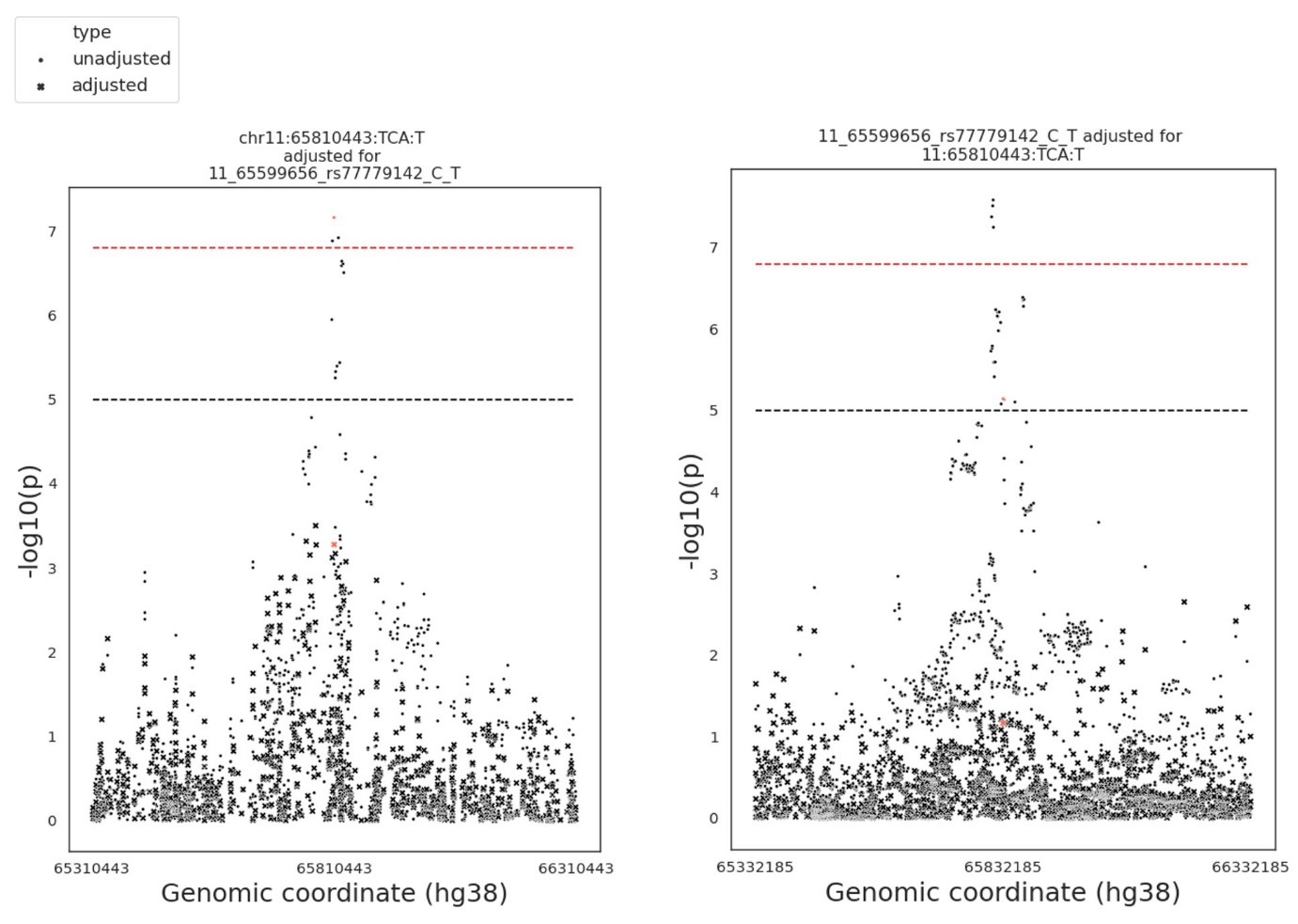

**Supplementary Figure 11.** STR-SNP pair: chr11:65810443:TCA:T- rs77779142.

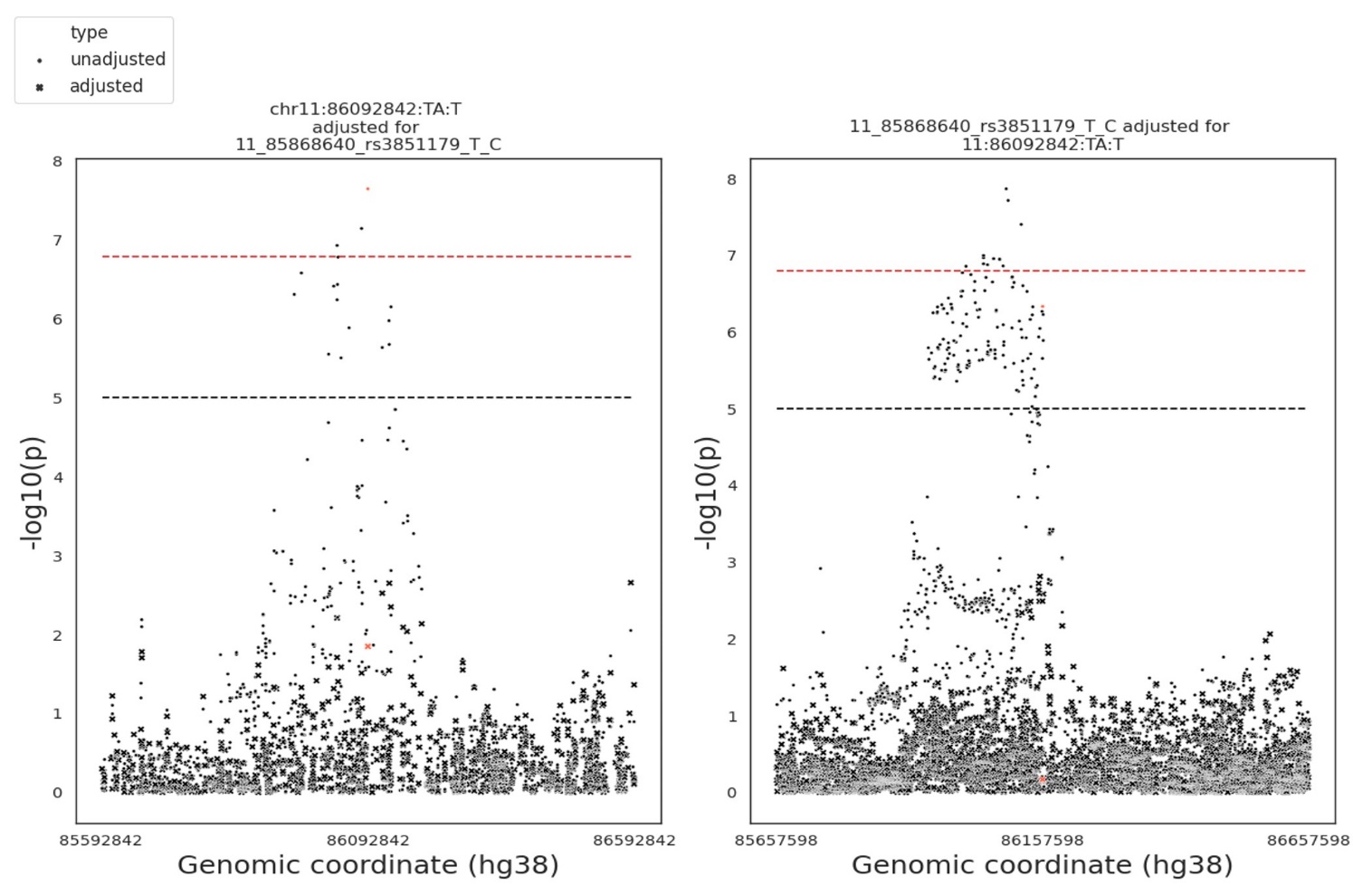

**Supplementary Figure 12.** STR-SNP pair: chr11:86092842:TA:T - rs3851179.

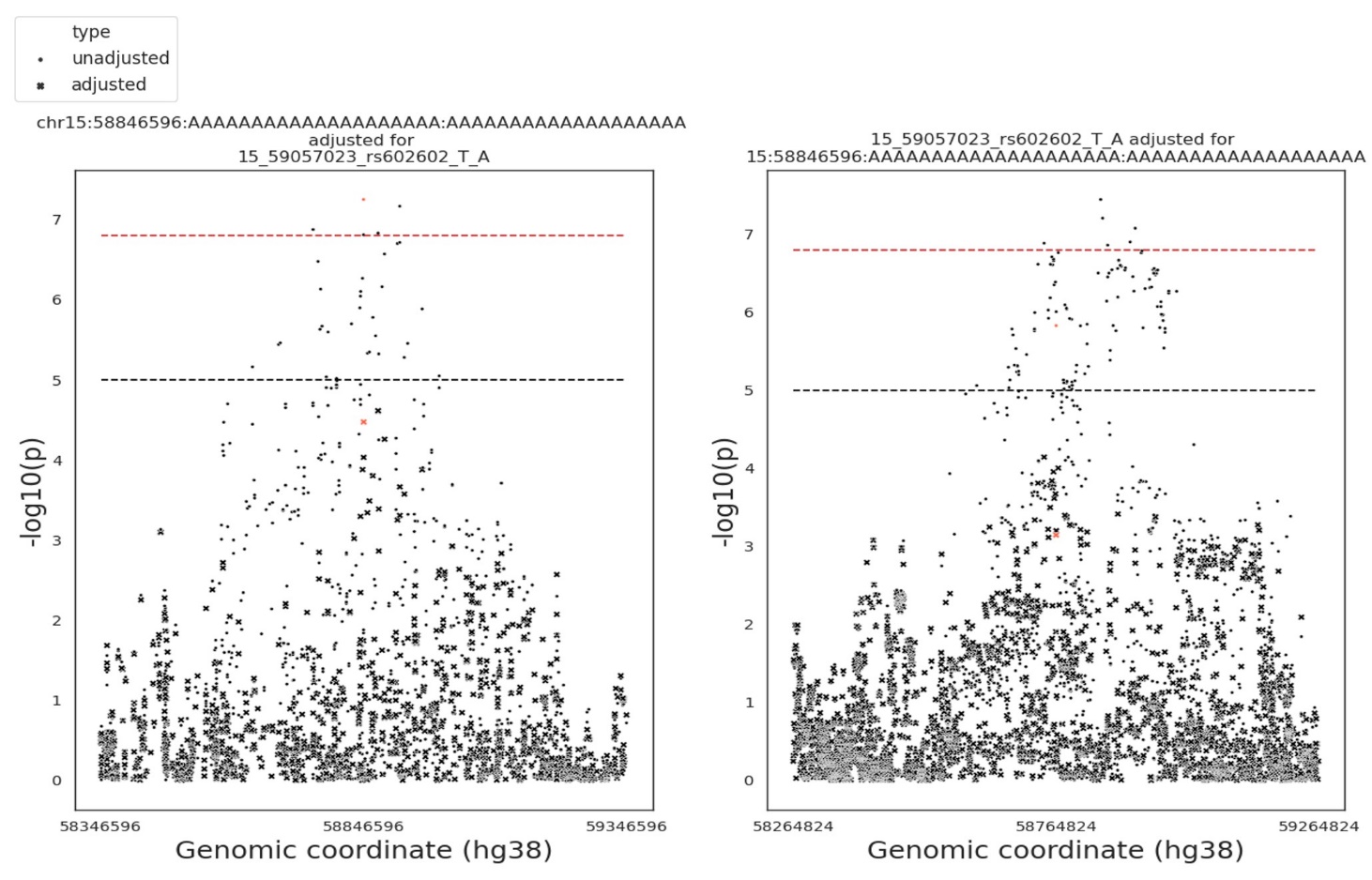

**Supplementary Figure 13.** STR-SNP pair: chr15:58846596:A20:A19- rs602602.

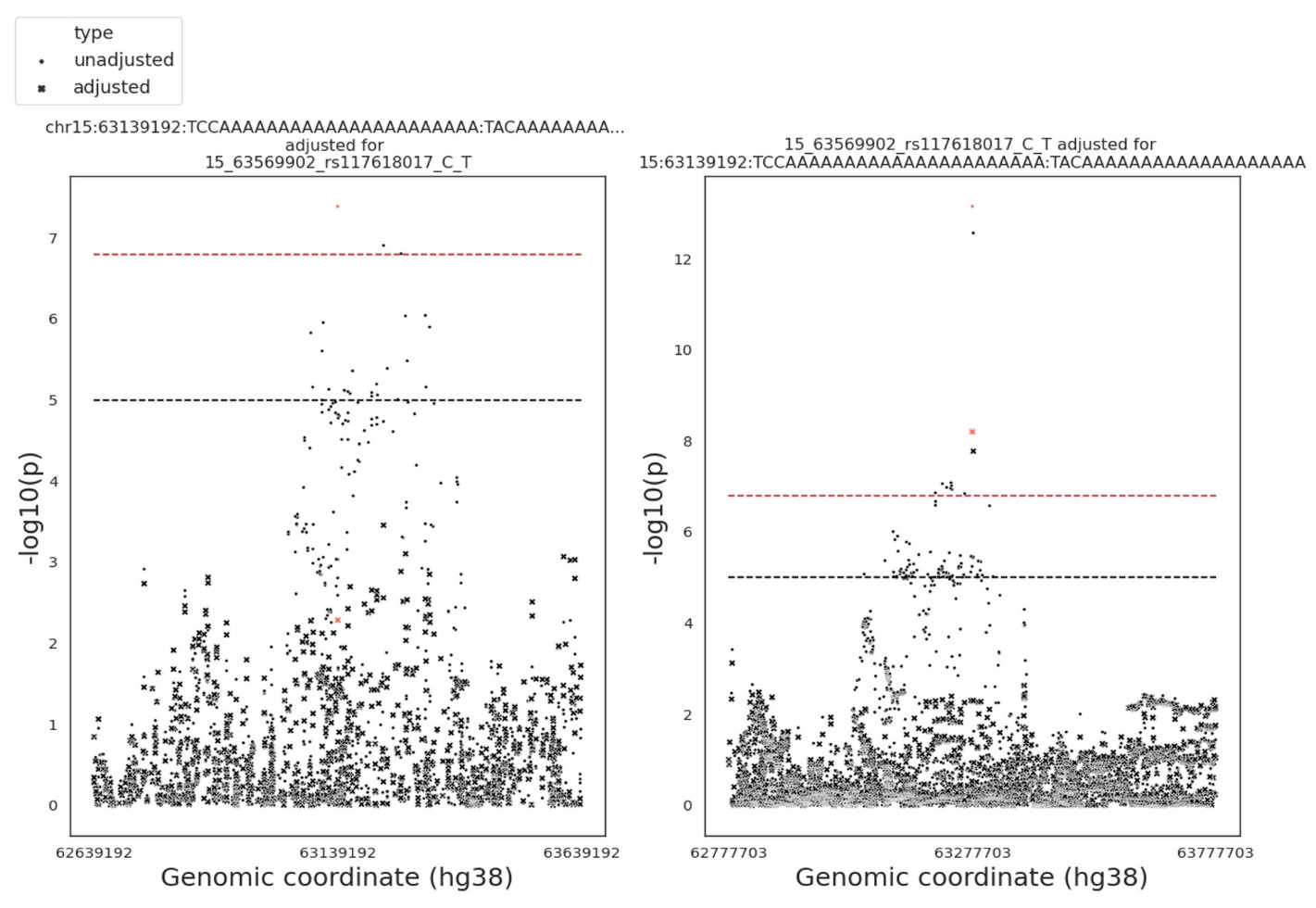

**Supplementary Figure 14.** STR-SNP pair: chr15:63139192:TCC(A)22:TAC(A)19 - rs117618017.

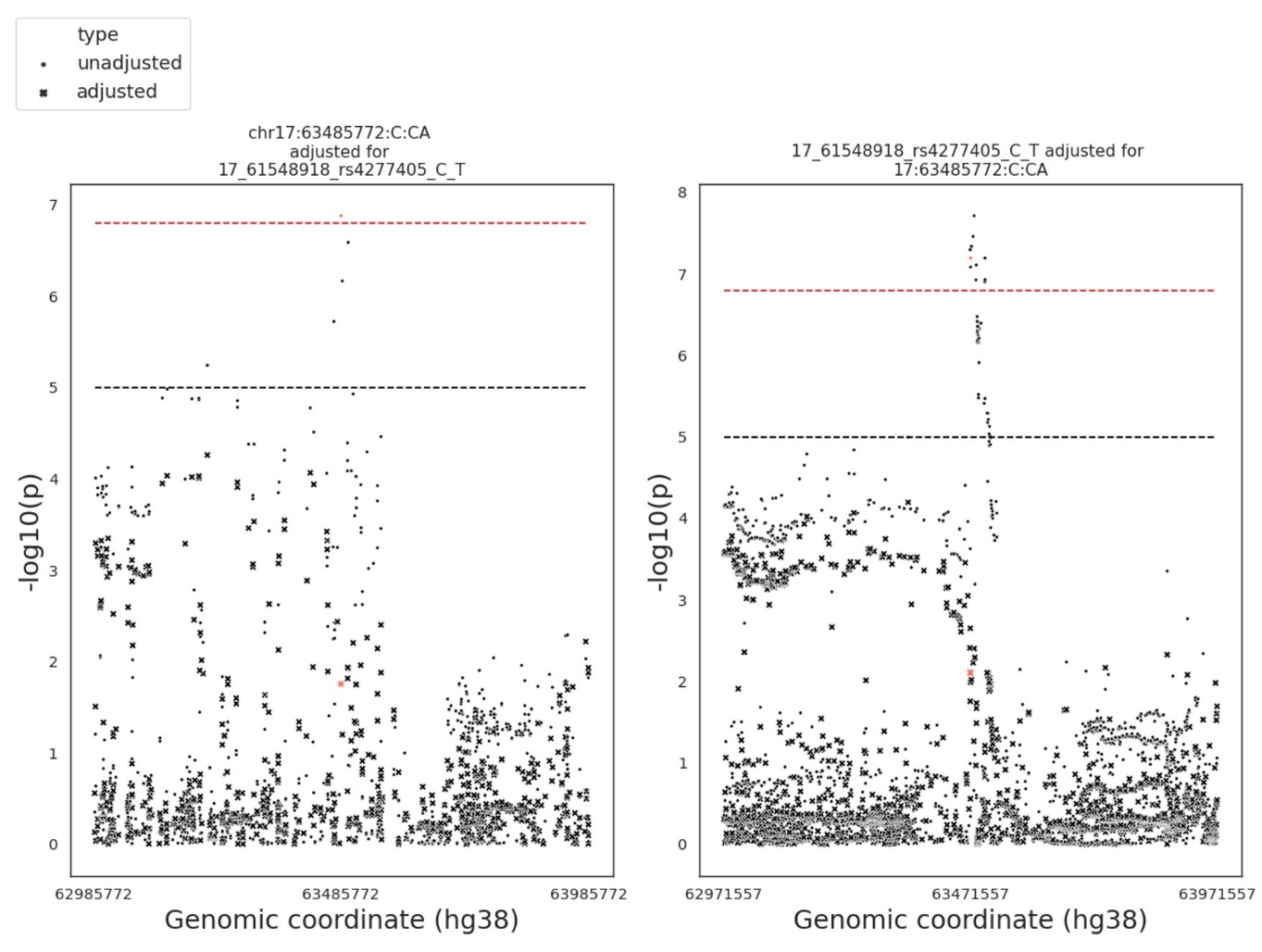

**Supplementary Figure 15.** STR-SNP pair: chr17:63485772:C:CA - rs4277405.

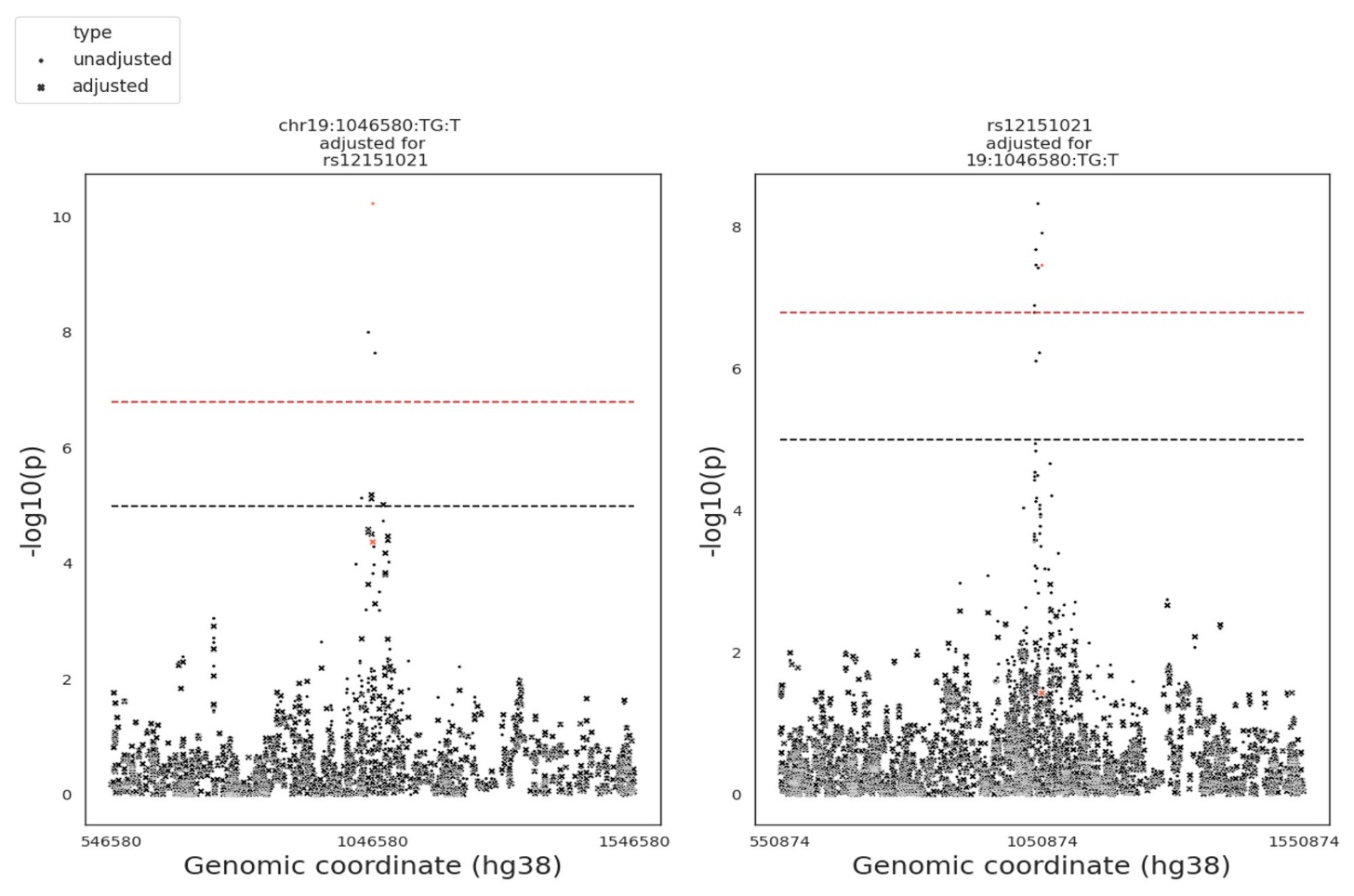

**Supplementary Figure 16.** STR-SNP pair: chr19:1046580:TG:T - rs12151021.

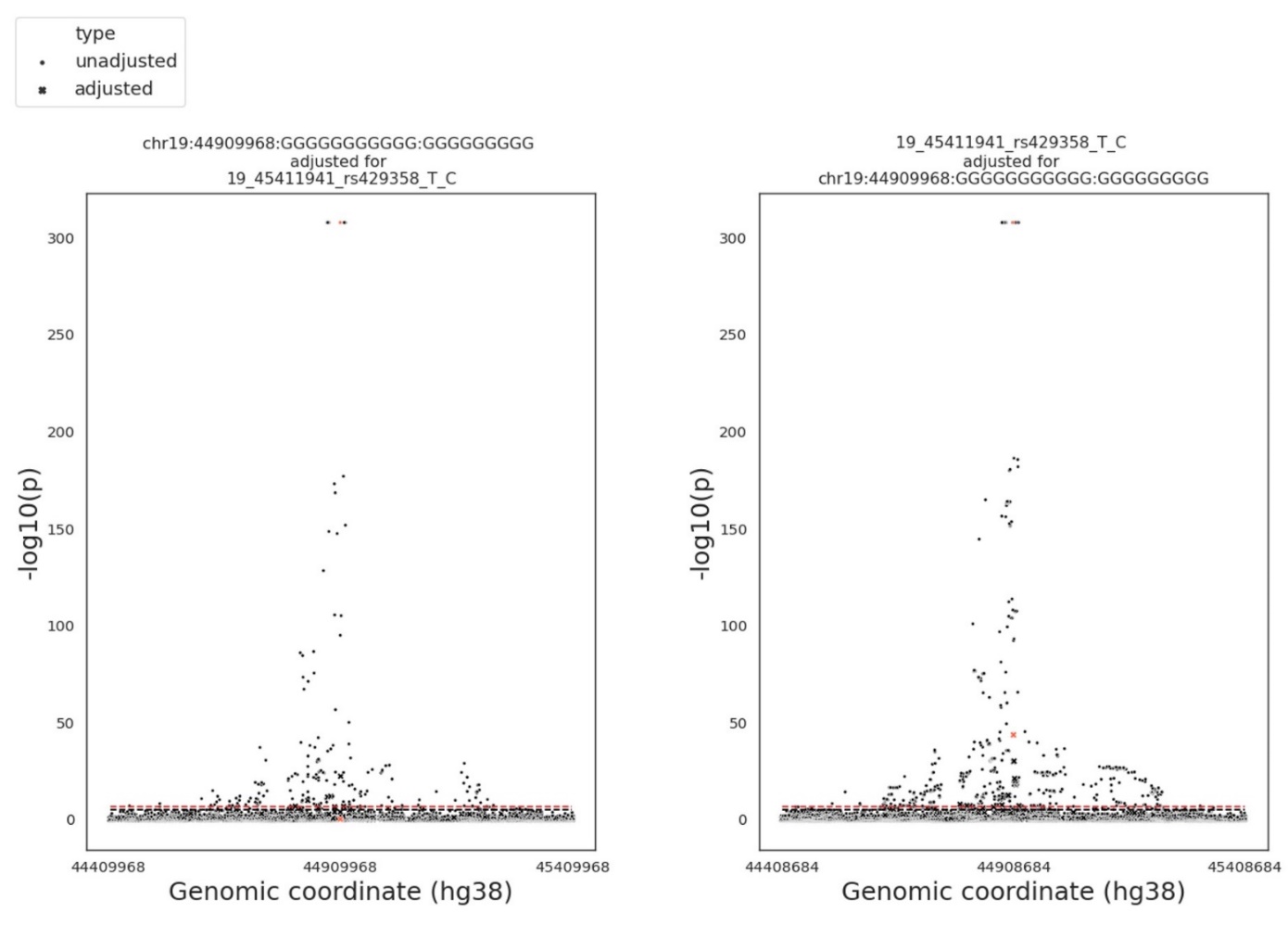

**Supplementary Figure 17.** STR-SNP pair: chr19:44909968:G11:G9 - rs429358.

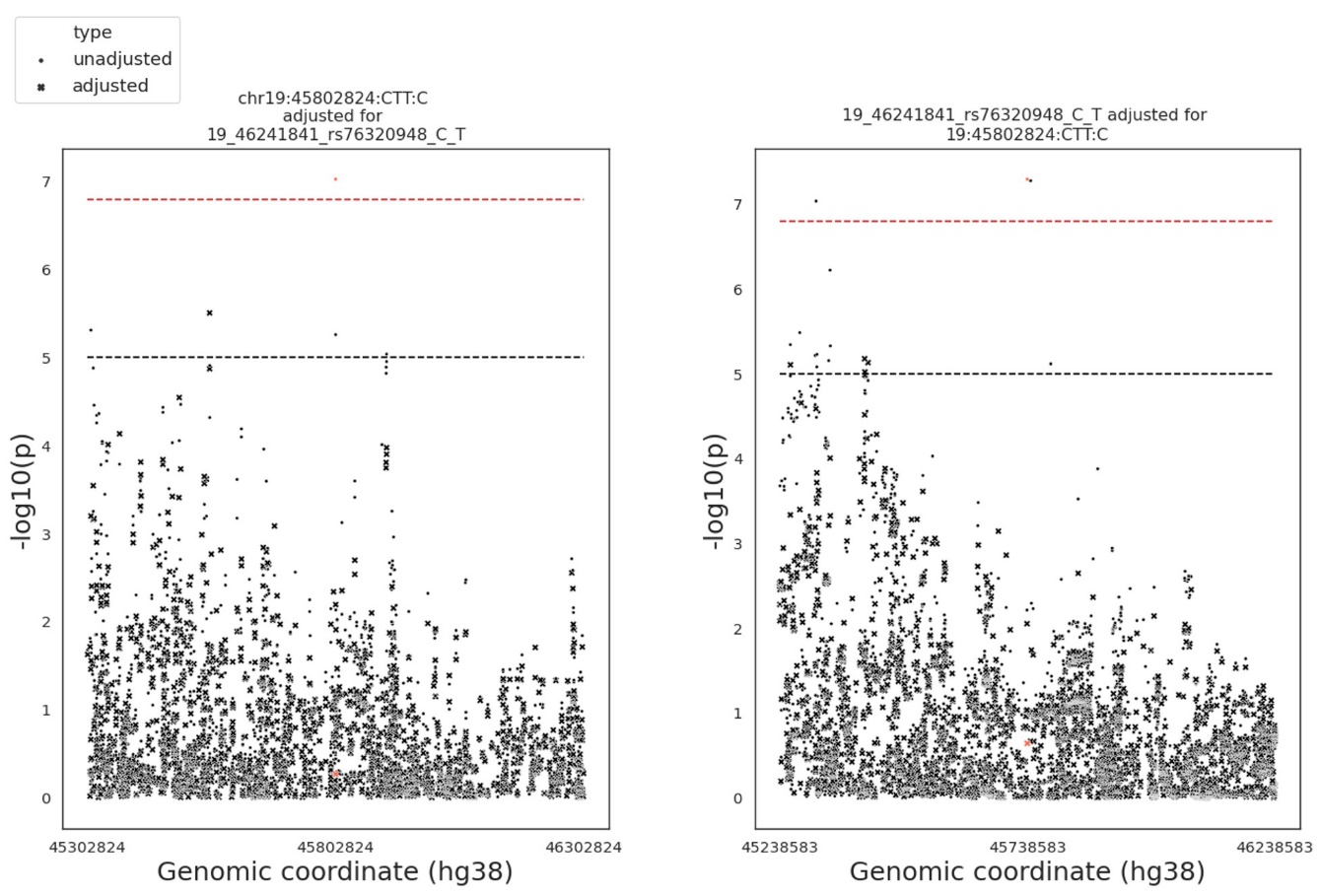

**Supplementary Figure 18.** STR-SNP pair: chr19:45802824:CTT:C - rs76320948

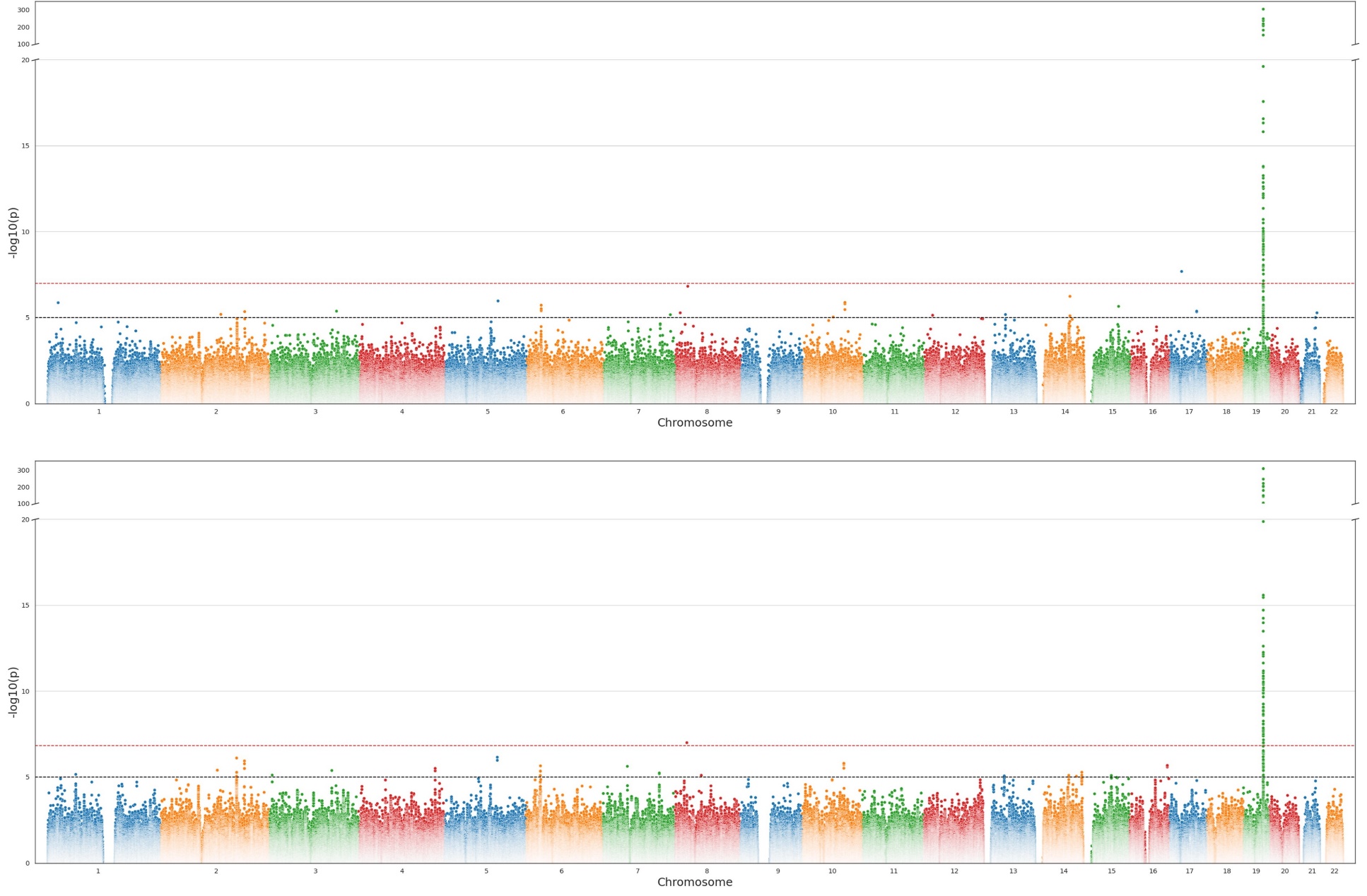

**Supplementary Figure 19.** Comparison of GWAS results performed on “White-British” subset of samples with WGS data available (n = 95,201) using genotyped STRs (upper panel) and imputed STRs (lower panel).

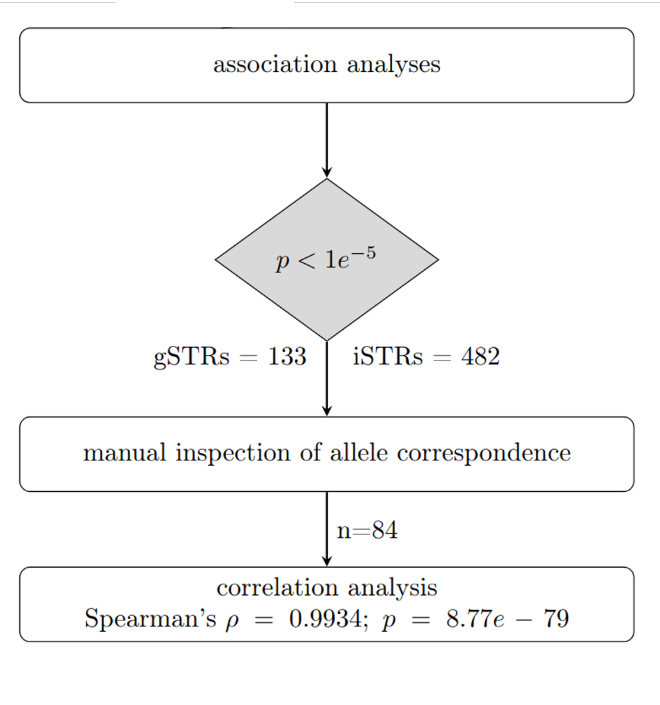

**Supplementary Figure 20.** Flowchart of matching procedure between imputed and WGS-derived STRs.

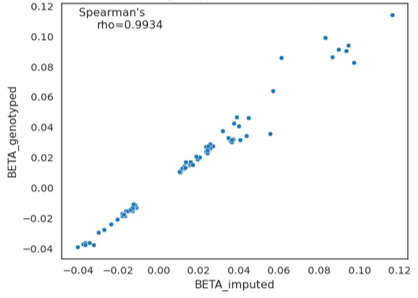

**Supplementary Figure 21.** Correlation plots of GWAS effect sizes (beta).

STR variants with p<1.0E-05 in either imputed or genotyped dataset, manually curated matches (n=84; Methods).
